# Modelling the Effects of Smoking Behavior on Male-to-Male HPV Transmission and Anal Cancer Progression

**DOI:** 10.64898/2026.08.11.26360159

**Authors:** Rashidat Oluwabukola Owolabi, Maia Martcheva, Indrajit Ghosh

**Affiliations:** Bagchi School of Public Health, Ahmedabad University, Ahmedabad, Gujarat, India; Department of Mathematics, University of Florida, Gainesville, Florida, United States of America

**Keywords:** Human papillomaviruses, Anal cancer, Mathematical model, Backward bifurcation, Global sensitivity analysis, Computational simulation

## Abstract

Human Papillomavirus (HPV) infection among men who have sex with men (MSM) has become a significant public health concern, particularly in countries where male vaccination is unavailable. Given the high susceptibility of MSM to HPV and anal cancer, and the unavailability of HPV vaccination for males in low- and middle-income countries (LMICs), there is a need to identify alternative interventions for reducing disease transmission and burden in this population. The novel mathematical model presented in this article couples smoking behavior dynamics with HPV transmission and anal cancer progression among MSM. Smoking reduction is introduced as an intervention to assess its effects on disease transmission and burden. The basic reproduction number (*R*_0_) is derived using the next-generation matrix method, and a global sensitivity analysis is performed using partial rank correlation coefficients (PRCC) to identify the influence of model parameters on *R*_0_. Further, the theoretical analysis of the model reveals a backward bifurcation, implying that *R*_0_ < 1 is necessary but not sufficient to eradicate the disease. The study finds that smoking reduction among MSM reduces HPV infection and anal cancer burden relative to baseline projections without intervention. The joint effect of smoking reduction and vaccination shows that the critical vaccination coverage needed to achieve *R*_0_ < 1 decreases as the level of smoking reduction increases. A similar outcome is observed for contact reduction. These findings highlight the importance of concurrent interventions, which can significantly curtail the spread of HPV and reduce disease burden in both the high-risk group and the general population.

## 1 Introduction

Human papillomaviruses (HPVs) are transmitted sexually and are tropic for cutaneous and mucosal cells. Although the immune system typically eradicates the virus, persistent infections can lead to cancer. More than 5% and 30% of all cancers and cancers of infectious etiology, respectively, are HPV-associated [1]. HPV has been identified to have a significant impact on men, causing diseases substantially similar to those in females. The precursor lesions and cancers of the anus, genitals, and oropharynx in men are now linked to HPV [2]. The natural history of HPV has been described in terms of the detection of new infection or reinfection, and as an undetected virus due to its absence from the body. The updated natural history differs somewhat, encompassing auto-inoculation, acquisition from recent sexual activity, or detection of a dormant infection; moreover, immune control can render the virus undetectable rather than achieving complete viral clearance [3]. High-risk HPV infections that persist over a prolonged period can lead to the development of cancer in the infected part of the body. This virus infects the squamous cells lining the surfaces of organs [4].

HPV infection is transmitted primarily through sexual contact of various forms, including the use of sexual devices. The body’s immune system can clear the infection in most cases, but it remains imperative to develop strategies to reduce transmission and cancer progression. Such preventive strategies include condom use, smoking cessation, and circumcision, especially among heterosexual couples [5]. Most infections clear naturally within two years, even as the incidence of infection increases. While some infections are asymptomatic and do not progress to clinical disease, they may persist in a small proportion of infected individuals. Persistent infection has been implicated as a major risk factor for progression to cervical cancer, a pathogenetic pathway analogous to that of other HPV-related cancers. The virus can be transmitted during both the acute and persistent phases of infection [6].

The substantial impact of HPV-related cancers in men has been the focus of recent studies, as previous research paid greater attention to the effects of HPV on females [5]. Among males aged 15 years and above, genital HPV can be found in 1 in 3 individuals, and 1 in 5 males is infected with one or more high-risk or oncogenic HPV types, indicating that genital HPV infections are commonly harbored by men. It is therefore imperative to design preventive and control strategies for HPV infection in men to reduce the overall burden of infection and associated diseases in both males and females [7]. HPV has been implicated in the global rise in the incidence of anal carcinoma (AC), particularly among men who have sex with men (MSM) [8]. This increase in AC incidence is primarily attributable to the increased prevalence of HPV. While incidence is currently highest in high-income countries, it is expected to rise in low-income countries (LICs) due to higher HPV prevalence and lower vaccine coverage [9].

HPV infection is a significant public health concern. Compared to women and heterosexual men, MSM have a higher susceptibility to HPV [10]. The incidence, persistent infection, and clearance rates of any anal HPV infection among MSM are 43.6, 23.4, and 58.3 per 1,000 person-months, respectively [11]. Although the median duration of any anal HPV and high-risk HPV (HR-HPV) infection has been reported as 9.67 and 8.51 months, with clearance rates of 50.9 and 62.1 per 1,000 person-months, respectively [10], naturally induced HPV antibodies do not confer protection against subsequent anal infection within one year among MSM [10, 12]. In Nigeria, there is a prevalence of 92% and 74% of any HPV and high-risk HR-HPV, respectively [13], and a prevalence of 91.1% and 40.6% of anal HR-HPV among HIV^+^ and HIV^*−*^ MSM, respectively [14]. At higher risk of HPV infection is the HIV^+^ MSM population. HIV^+^ MSM have a higher prevalence of anal HR-HPV, multiple anal HR-HPV types, and HSIL compared to HIV^*−*^ MSM [14, 13]. The incidence rate of anal cancer among HIV-positive MSM is substantially higher than among HIV-negative MSM (85 vs. 19 per 100,000 person-years) [15]. Rates of tobacco use, particularly cigarette smoking, are higher among MSM than in the general population [16]. The incidence, prevalence, and persistence of HPV are higher among current smokers than non-smokers [17]. It has been reported that smoking reduction is likely to increase the resolution of HPV infection compared to frequent, long-term, or heavy smoking [18]. Since vaccination is not available to protect susceptible males in Nigeria from acquiring HPV, the incidence and prevalence of HPV and AC among MSM may increase in the absence of alternative interventions. Smoking reduction could therefore play an important role in interrupting the transmission, acquisition, and burden of HPV and AC among MSM.

Several mathematical models have been used to describe the impact of various interventions on the transmission dynamics of HPV and cancer progression. These include assessments of: the effectiveness of vaccination and Pap screening [19]; two-dose vaccination [20]; changes in sexual behavior and smoking patterns in the female population [21]; how vaccination against HPV 16/18 can reduce the burden of HPV 31/45 [22]; the synergistic effects of awareness, screening, and vaccination [23, 24]; and the treatment of HPV and cervical cancer [25]. The impact of vaccination was also evaluated in a study combining heterosexual males and females with the MSM population, which found that MSM derive lower benefits from vaccination of the heterosexual population and should therefore be given higher priority in vaccination programs [26].

This study considers low- and middle-income countries where HPV vaccination is available only to the female population. The novelty of this model lies in the explicit coupling of smoking behavior dynamics with transmission and disease progression, and in the evaluation of smoking reduction as an alternative intervention to curtail HPV transmission and anal cancer progression in a high-risk group, addressing gaps in previous studies that mostly focused on vaccination of the male population. The main objective of this study is to determine whether smoking reduction has a substantial impact on the transmission, incidence, and burden of HPV and anal cancer in the MSM population.

## 2 Model Construction

This model has eight compartments for the total population *N*_*m*_(*t*). The description of the compartments are presented in Table 1. The *S*_*n*_(*t*) is the susceptible non-smokers, *S*_*s*_(*t*) is the susceptible smokers, *E*(*t*) is the exposed individuals, *I*_*a*_(*t*) is the individuals with acute infection, *I*_*p*_(*t*) is the individuals with persistent infection, *P*_*c*_(*t*) is the individuals with High Grade Squamous Interepithelial lesion (HSIL), *A*_*C*_(*t*) is the individuals with anal cancer, and *R*_*HPV*_ (*t*) is the individuals who have recovered from HPV infection. Therefore,

**Table 1:** This table defines the eight mutually exclusive epidemiological compartments partitioning the total MSM population.

| State Variables | Description |
| --- | --- |
| $S_n$ | Number of susceptible males who are non-smokers |
| $S_s$ | Number of susceptible males who are smokers |
| $E$ | Number of males who are exposed to HPV infection |
| $I_a$ | Number of males who have an acute infection |
| $I_p$ | Number of males who have a persistent infection |
| $P_c$ | Number of males who have HSIL |
| $A_C$ | Number of males who have anal carcinoma |
| $R_{HPV}$ | Number of males who recovered from HPV infection |

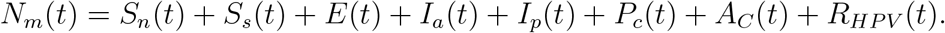

The force of infection *λ*_*mm*_ is given by

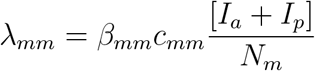

where *β*_*mm*_ is the probability of HPV transmission from infectious males to susceptible males. *c*_*mm*_ is the average number of sexual partners a sexually active male has with males.

### 2.1 Model Assumptions

- The susceptible population consists of men 15 years and above.
- There is a homogeneous mixture of individuals in the population, i.e., every sexually active man is equally likely to mix and have homosexual contact with other sexually active males.
- Non-smokers have a reduction in HPV acquisition.
- Infected individuals are infectious before virus clearance.
- There is no prior immunity from vaccination.
- Immunity derived from infection wanes, and recovery from HPV infection can return to either of the susceptible classes.
- In LMICs, anal cancer screening, detection, and treatment are poor; therefore, individuals in the *A*_*C*_ compartments are assumed to be undetected and untreated.
- All model parameters are positive.

Based on the flow chart in Figure 1, the following system of normal differential equations is derived:

**Figure 1:**
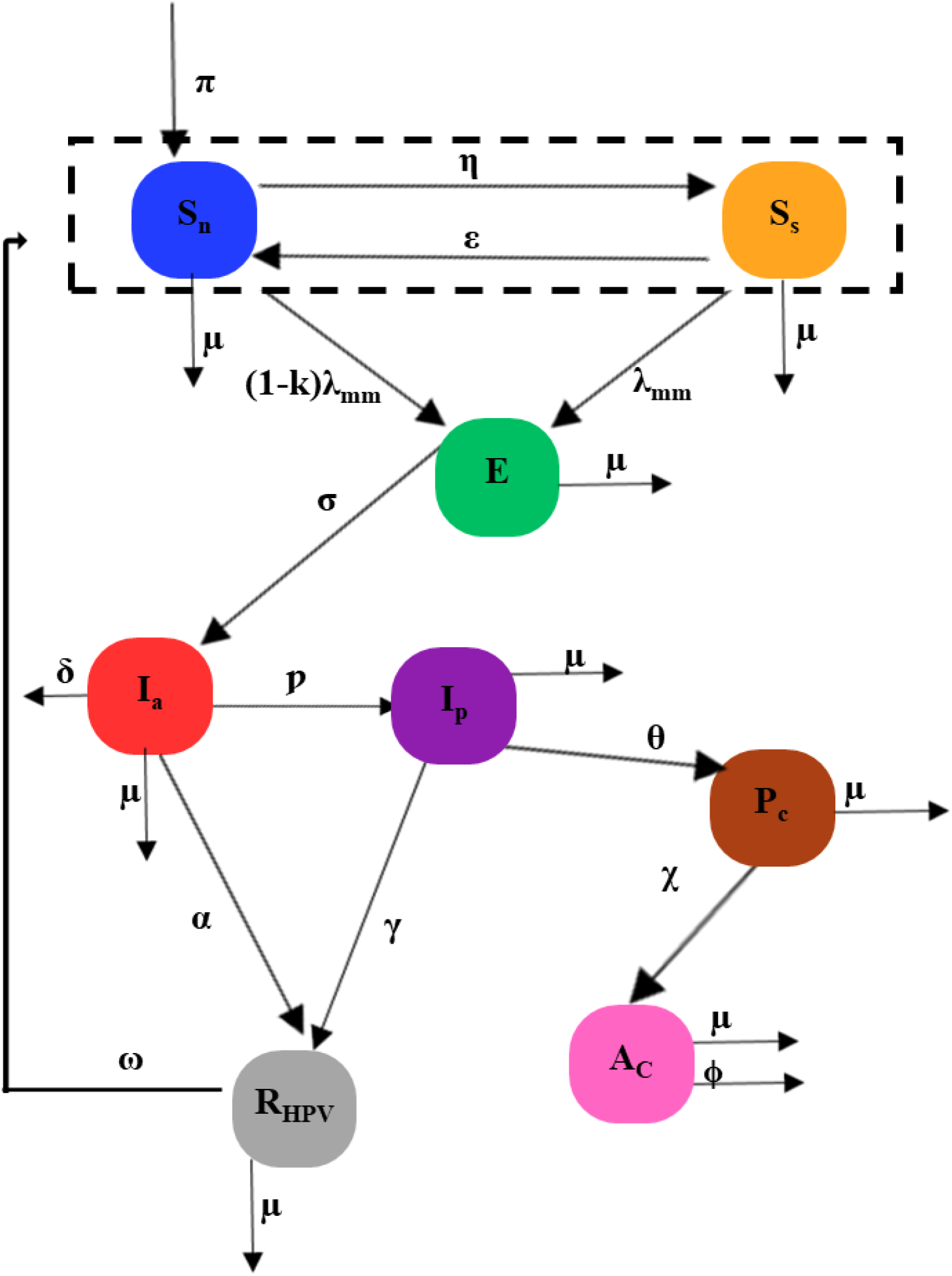
Schematic diagram of Male-to-Male HPV transmission and anal cancer progression. This compartmental framework maps the flow of the MSM population across varying stages of smoking behavior and disease states. The model uniquely couples smoking initiation and cessation pathways with the biological progression of HPV, from acquisition through to anal carcinoma, providing a comprehensive representation of overlapping risk factors.

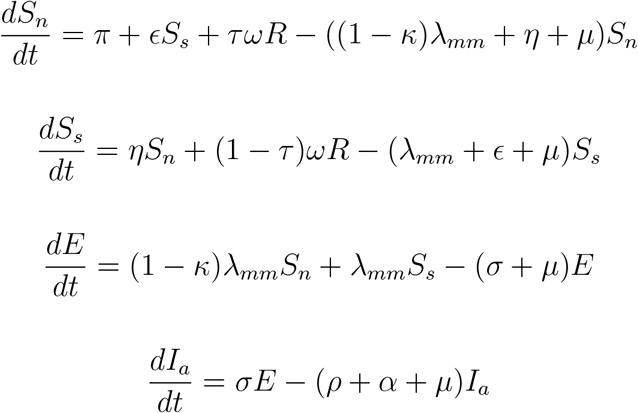

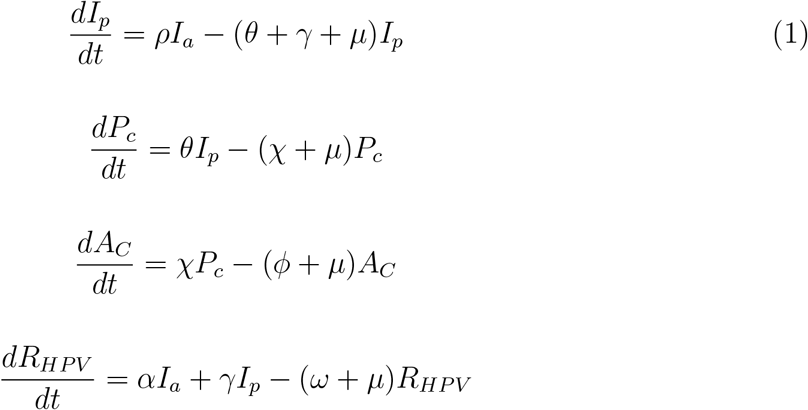

The susceptible non-smoker compartment increases with the recruitment rate *π*, the rate at which smokers quit *ϵ*, and the rate at which waning immunity from the recovery class occurs *τω* (*τ* is the proportion of the recovery class that returns to the non-smoker compartment). Breakthrough infection occurs at a rate (1*− κ*)*λ*_*mm*_, where 0 *≤k ≤*1 is the efficacy of non-smoking status in reducing the risk of HPV acquisition. The class is further reduced by non-smokers who transition to smokers at a rate *η*. Individuals in all compartments are assumed to die naturally at a rate *µ*. The susceptible smoker compartment is further increased by the waning of immunity from the recovery class at the rate (1 *− τ*)*ω*, and is reduced by HPV acquisition at the rate *λ*_*mm*_. Progression from exposed to acute infection occurs at a rate *σ*. There is a reduction in the acute-infection class, with individuals progressing to persistent infection at a rate *ρ* and recovering at a rate *α*. The progression from persistent infection to High Grade Squamous Intraepithelial Lesion (HSIL) occurs at a rate *θ*, and further reduction in the compartment is due to recovery at a rate *γ*. The transition out of the HSIL class occurs at a rate *χ* to the Anal carcinoma class, and death due to AC among the MSM occurs at a rate *ϕ*.

## 3 Mathematical Analysis

### 3.1 Disease-Free Equilibrium and Basic Reproduction Number

The disease-free equilibrium (DFE), denoted by 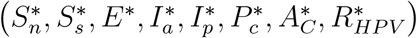, is obtained as:

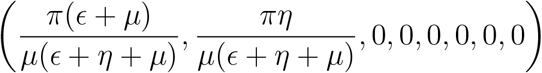

The infected compartments are *x* = [*E*(*t*), *I*_*a*_(*t*), *I*_*p*_(*t*)]^*T*^ . They can be written as follows:

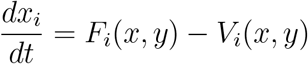

Where *F*_*i*_ is the rate of appearance of new infections, and *V*_*i*_ represents the remaining transitional terms (births, deaths, progression, recovery).

At DFE, we find that the matrices *F* and *V* are given by:

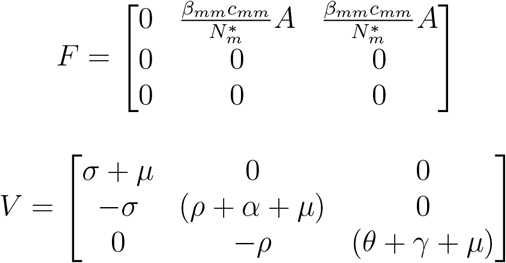

Therefore,

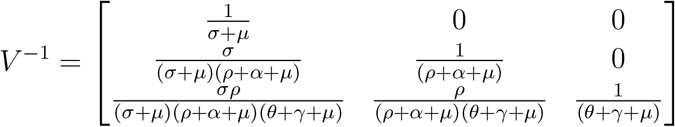

where 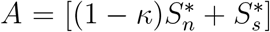. Thus, the Next-Generation Matrix is given by *K* = *FV* ^*−*1^ [37]:

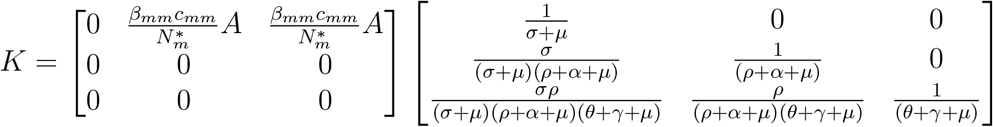

The basic reproduction number, *R*_0_, is the spectral radius of the matrix *K*. Since the matrix *K* only has non-zero entries in its first row, its spectral radius is simply the trace (the *K*_11_ element). Factoring out the common terms correctly yields:

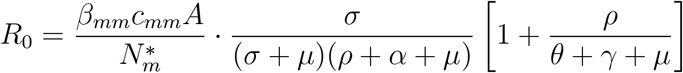

Substituting the equilibrium values 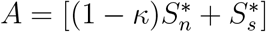 and 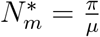

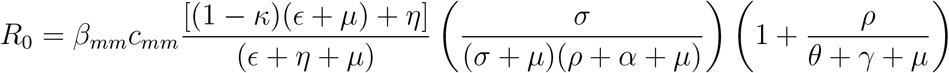

In epidemiology, *R*_0_ represents the expected number of secondary infections produced by a single infected individual over their entire infectious period when introduced into a completely susceptible population.

By expanding and grouping the terms in the final equation, we can interpret *R*_0_ as the product of four distinct epidemiological factors:

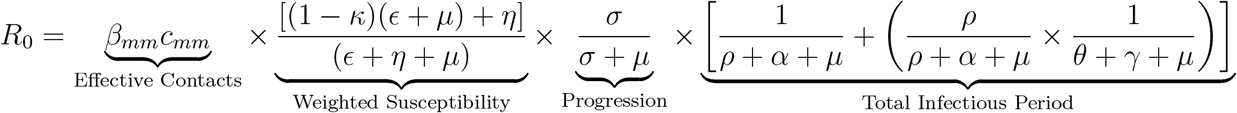

The effective contact rates: This is the baseline rate at which an infectious individual spreads the virus. It is the product of the probability of HPV transmission per sexual partnership (*β*_*mm*_) and the average number of sexual partners a sexually active male has with other males per unit of time (*c*_*mm*_).

The weighted susceptible pool (effect of smoking): This represents the effective proportion of the population that is susceptible to acquiring the infection at the DFE. It is essentially the term 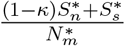 . Because non-smokers have a reduced risk of HPV acquisition (efficacy *κ*), this term mathematically penalizes the baseline transmission rate based on the ratio of non-smokers to smokers in the population. It is governed by the natural death rate (*µ*) and the continuous behavioral transitions of quitting smoking (*ϵ*) and initiating smoking (*η*).

Progression probability: When an individual is newly exposed to HPV, they enter the latent/exposed compartment (*E*). This fraction is the probability that the individual successfully progresses to the acute infectious stage (*σ*) rather than dying from natural, non-disease-related causes (*µ*) during the latency period.

The total effective infectious period: As the model assumes individuals are infectious during both the acute and persistent phases, the total infectious period is the sum of the time spent transmitting the virus in both compartments.

Acute Phase Duration 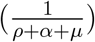: This is the average duration an individual spends in the acute infection compartment before either progressing to a persistent infection (*ρ*), recovering (*α*), or dying naturally (*µ*).

Persistent Phase Duration 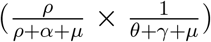: To transmit during the persistent phase, an individual must first survive the acute phase and progress to the persistent phase (a probability of 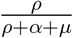). If they do, they spend an average time of 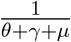 in the persistent state before progressing to precancer/HSIL (*θ*), recovering (*γ*), or dying (*µ*).

In short, the expression of *R*_0_ reflects the sequence of the disease: (Rate of contacts) *×* (Probability the contact is susceptible given smoking status) *×* (Probability of surviving latency) *×* (Total time spent in acute and persistent infectious states).

### 3.2 Endemic Equilibria and Backward Bifurcation

Endemic equilibria are solutions to the system

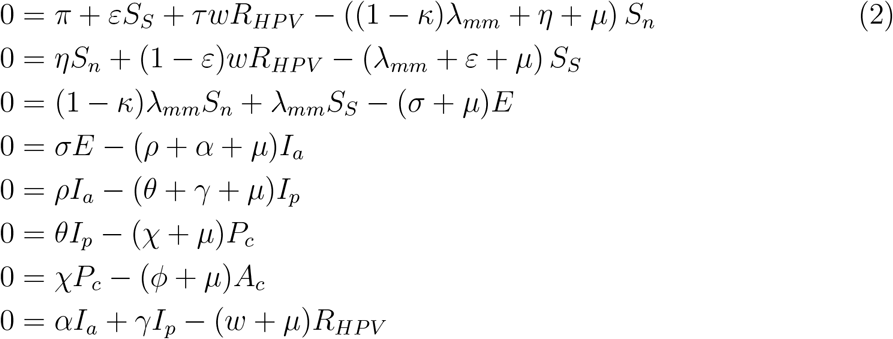

where

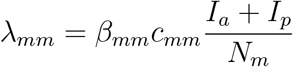

The equation for the total population size is

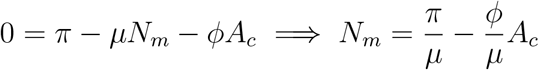

Solving for *I*_*a*_ we get

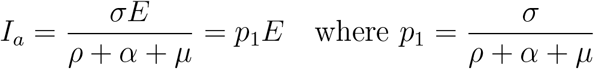

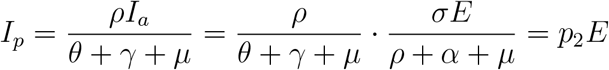

where 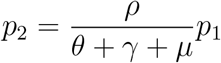

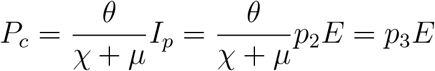

where 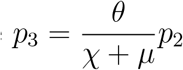

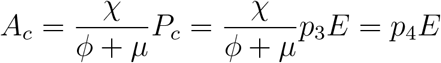

where 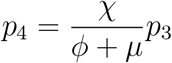. Finally

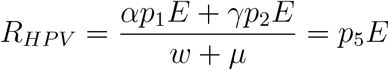

where 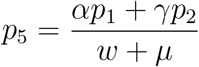

Solving the first two equations for *S*_*s*_ and *S*_*n*_ we get:

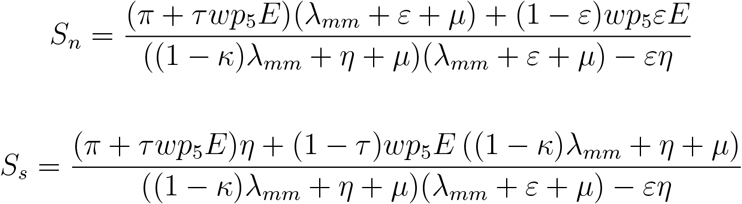

We note that

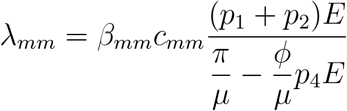

From the equation for *E* we have the following equation in *E*:

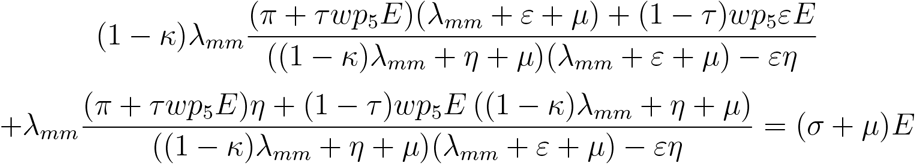

Since *E ≠* 0, we can cancel *E* from *λ*_*mm*_ and the right-hand side. Replacing *λ*_*mm*_ with its expression, and taking a common denominator we have the following equation in *E*:

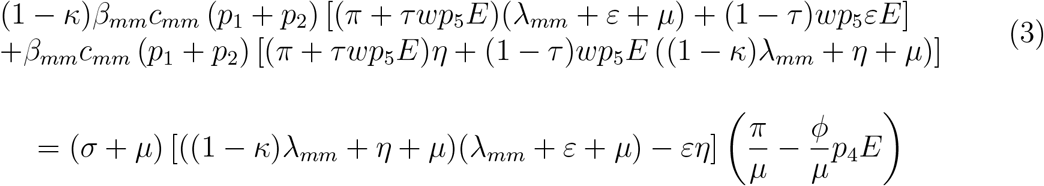

Denoting the LHS of this equation by *f* (*E*) and the RHS by *g*(*E*), this equation can be written as *f* (*E*) = *g*(*E*). Assume *R*_0_ *>* 1, then

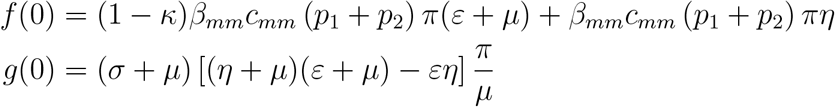

Thus, if *R*_0_ *≥* 1, then *f* (0) *≥ g*(0).

*f* (*E*) and *g*(*E*) are continuous functions of *E*. Multiplying both sides of *f* (*E*) = *g*(*E*) by 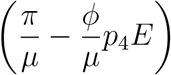 and taking the limit as 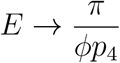 (denote 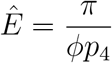) we get:

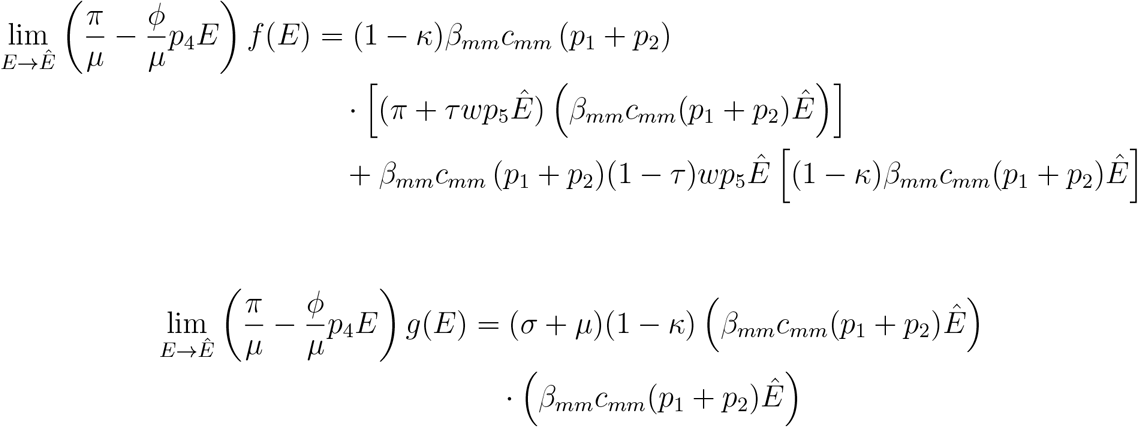

Dividing both sides by (*β*_*mm*_*c*_*mm*_)^2^, (1 *− κ*), *Ê*2, (*p*_1_ + *p*_2_)^2^ we have to compare

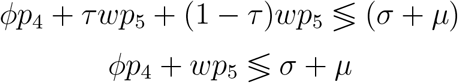

Consider

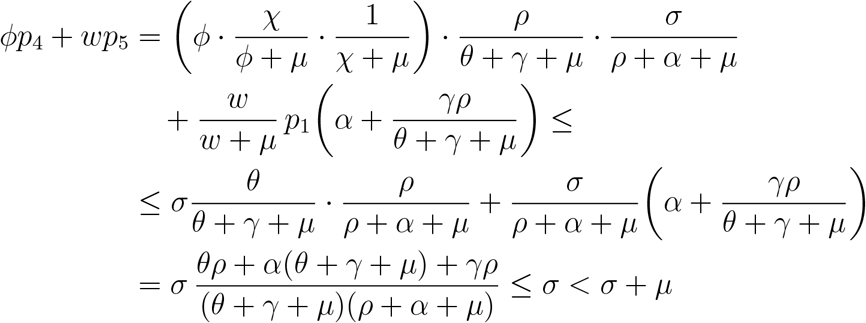

Thus, if *R*_0_ *>* 1, then the equation *f* (*E*) = *g*(*E*) has at least one solution *E*^*∗*^ *>* 0 and at least one endemic equilibrium exists. Assume *R*_0_ < 1. Then, we find a condition for backward bifurcation. To determine a condition for backward bifurcation, we consider (3) and think that *E* is a function of *β*_*mm*_. We differentiate (3) w.r.t. *β*_*mm*_. Then we set 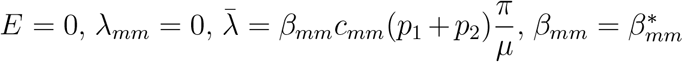 where 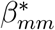 is the value that makes 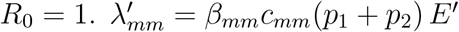

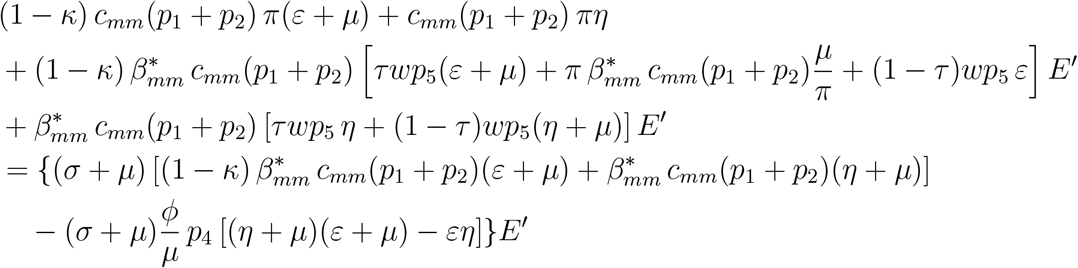

where

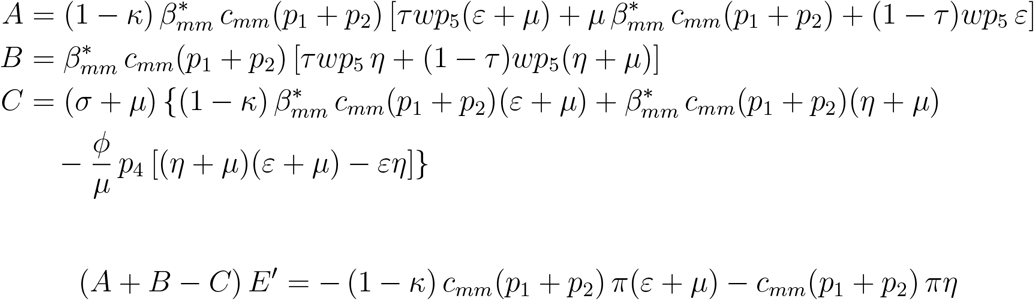

Thus, the condition for backward bifurcation is

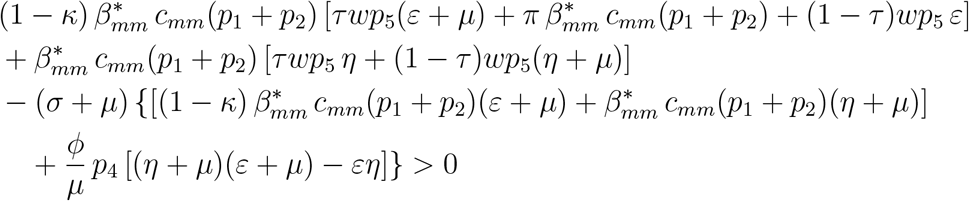

Dividing by 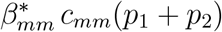 we have

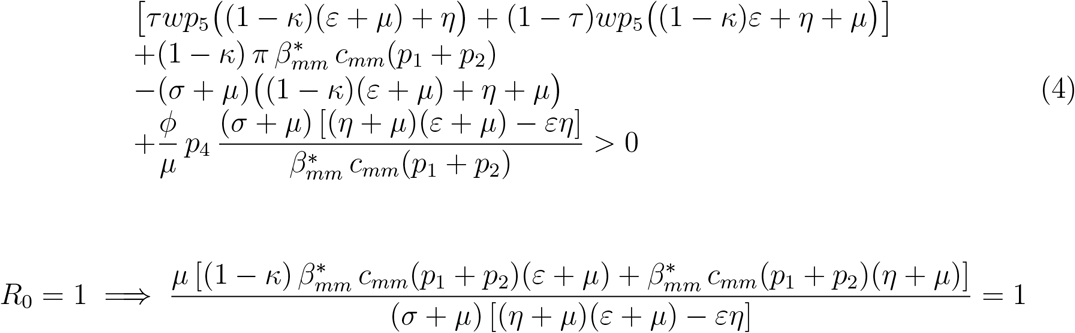

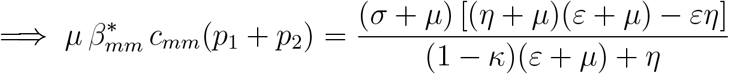

Thus, equation (4) becomes

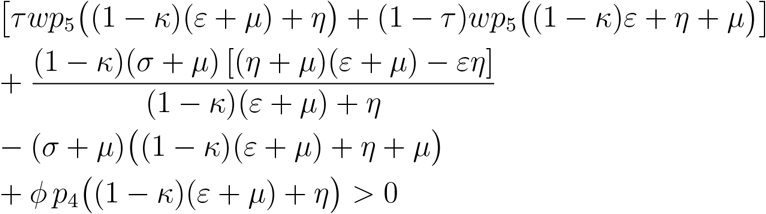

Thus, we have the following Theorem.

**Theorem:** If ℛ _0_ *>* 1, then system (1) has at least one endemic equilibrium. If ℛ_0_ < 1, then system (1) may have no endemic equilibria. Backward bifurcation and multiple equilibria exist if and only if

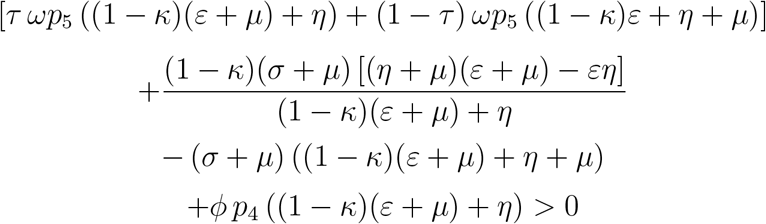

The backward bifurcation is illustrated in Figure 2.

**Figure 2:**
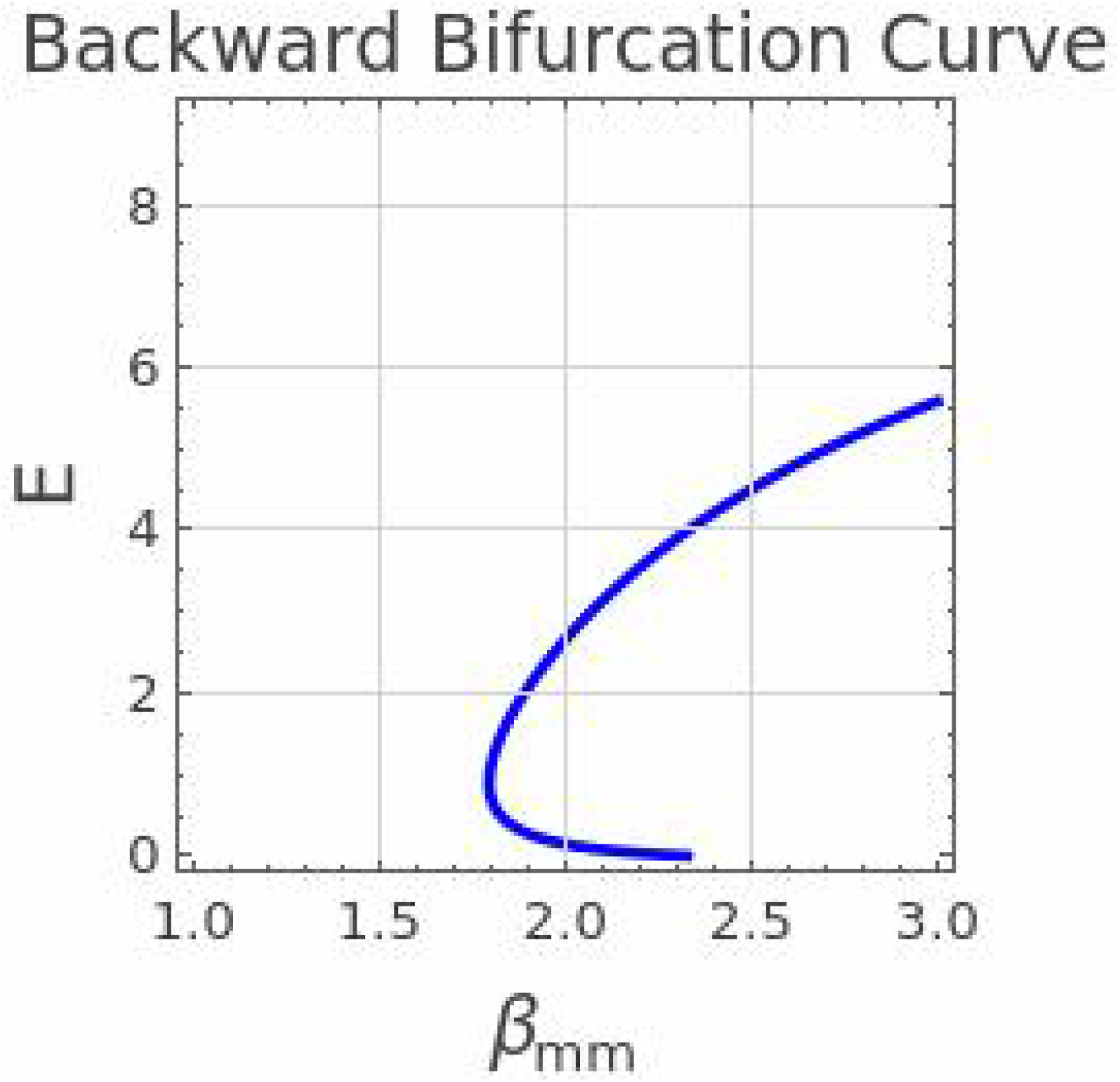
Backward bifurcation diagram for system (1). Parameter values: *σ* = 15, *ρ* = 0.4, *α* = 3, *µ* = 0.01, *θ* = 0.3, *γ* = 2, *χ* = 0.25, *ϕ* = 4, *ω* = 0.2, *ε* = 0.0, *η* = 0.0, *κ* = 0.75, *τ* = 0.5, *π* = 10.0, *c*_*mm*_ = 5.0.

From an epidemiological perspective, the presence of backward bifurcation phenomenon implies that the traditional condition for eliminating disease by lowering the basic reproduction number (*R*_0_) to below one (*R*_0_ < 1) is not adequate for ensuring the elimination of disease [38]. In the case of a backward bifurcation, even at *R*_0_ < 1, a stable disease-free equilibrium coexists with a stable endemic equilibrium. Therefore, if the prevalence level of HPV in the MSM community is initially very high, the disease will continue to be endemic even if public health measures manage to reduce *R*_0_ < 1. Effective public health measures have to be adopted in a way to either reduce *R*_0_ to a sub-threshold value or eliminate the initial infectious individuals as quickly as possible.

## 4 Computational Simulation

### 4.1 The State Variables

The value of the initial susceptible non-smoker population (*S*_*n*_) was estimated from Nigerian demographic data [28]. The initial male population was estimated by summing the population of males aged 15 years and older (10686788). The proportion of MSM in Nigeria was reported as 0.0126 [27]. The total population of MSM was estimated as *≈* 134654 (0.0126 *×* 10686788). One infected person was introduced to the population in the exposed (E) compartment, while all other compartments were set to zero at the initial state.

### 4.2 The Model Parameters

There is a scarcity of data on HPV and anal cancer among MSM in Nigeria; however, some data on HPV infection and anal cancer were derived from studies among MSM in Nigeria [14, 13]. Where local values were unavailable for the parameters, estimates were derived from global and regional studies among MSM. The model looks back to transmission from 1950; data are scarce on infection and disease progression around this period. Estimates were made using values from nearby years that were available and suitable for the model’s population. The parameters for smoking status transitions were estimated from a South African study, as no such data were available in Nigeria [30]. The model targets HIV uninfected MSM and uses estimates for this population where exclusively stated in the literature; otherwise, the general estimate was used. The summary of the parameter estimates and sources is presented in Table 2.

**Table 2:** This table details the demographic, behavioural, and biological parameters governing the system of ordinary differential equations. The baseline parameter values and their corresponding sources are mentioned.

| Parameters | Description | Baseline value | Source |
| --- | --- | --- | --- |
| $\pi$ | Recruitment rate of sexually active males | 5518 | Derived from [27, 28] |
| $\beta$ | Transmission rate from infected males to susceptible males | 0.5680 | Estimated from data [14] |
| $c$ | Average number of male sexual partners | 5 | Derived from [29] |
| $\eta$ | Transition rate from non-smoker to smoker | 0.073 | Derived from [30] |
| $\epsilon$ | Transition rate from smoker to non-smoker | 0.044 | Derived from [30] |
| $\mu$ | Natural death rate | 0.021 | Derived from [31] |
| $\omega$ | Rate of waning immunity | 0.8 | Derived from [12, 32] |
| $\tau$ | Proportion of recovery to non-smoker | 0.925 | Derived from [33] |
| $\kappa$ | Efficacy of non-smoking in reducing the acquisition of HPV infection | 0.423 | Derived from [17] |
| $\sigma$ | Progression rate from exposed to acute infection | 4 | Derived from [34] |
| $\rho$ | Progression rate from acute infection to persistent infection | 0.2472 | Derived from [11] |
| $\theta$ | Progression rate from persistent infection to HSIL | 0.0161 | Estimated from data [14] |
| $\chi$ | Progression rate from HSIL to anal cancer | 0.00432 | Estimated from data [15] |
| $\phi$ | Anal carcinoma-induced death rate | 0.034 | Derived from [35] |
| $\alpha$ | Recovery rate from acute infection | 0.66 | Derived from [11] |
| $\gamma$ | Recovery rate from persistent infection | 0.502 | Derived from [36] |

i. The recruitment rate (*π*) was estimated by assuming an equal proportion of males in the 10 – 14-year age bracket (2,189,678) [28].The population of 14-year-olds turning 15 (437935.6 ^*^ 0.0126 *≈* 5518) was used as the recruitment rate for the model, based on the assumption that susceptibles are 15 years and older.
ii. The average number of male partners (*c*_*mm*_) was obtained from a survey among MSM in Nigeria on the MSM sexual network, which reported a contact rate of 5 (3 – 10) anal sex male partners in a year [29].
iii. The transition from non-smoker to smoker and smoker to non-smoker, *η* and *ϵ*, respectively, were obtained from a South African longitudinal study with a 3-year follow-up time [30]. The study reported smoking initiation of 22.5%, relapse of 12.2%, and cessation of 12.3%. The individuals who initiated and relapsed were merged as non-smokers (19.7%) who transitioned to smokers at a rate of *η* = 0.073, and those who ceased smoking were estimated to transition to non-smokers at a rate of *ϵ* = 0.044 using the formula: 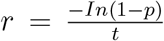, where r is the rate, p is the proportion, and t is the follow-up time.
iv. The natural death rate (*µ*) was estimated by using the life expectancy (47.14 years) of the year 2000,[31], since the model covers a 100-year span from 1950 – 2050 to reduce the differences in the life expectancy between these years. This results in a value of 0.021.
v. The waning rate (*ω*) was assumed to be 0.8. This is because studies have reported that natural antibodies from HPV infection do not offer protection against subsequent HPV infection among MSM [12, 32]. The model assumes that the susceptible population does not acquire immunity from vaccination.
vi. The proportion of recovered individuals that returned to the susceptible non-smokers compartment (*τ*) was estimated from the National Demographic Health Survey (NDHS) data [33]. The prevalence of tobacco use was 7.1% among men. The ratio of smokers to non-smokers was computed to be 0.08:1. This ratio was used to obtain the probability of non-smoking, which is 0.925 from 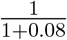.
vii. Current smokers are more likely to have the incidence of HR-HPV infection with an aOR =1.73 (95% CI: 1.00–2.99) [17]. The efficacy of non-smoking status (*κ*) to reduce the risk of acquisition of HPV infection was derived from this, with an estimated value of 0.423 from 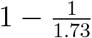.
viii. The latent period for HPV infection is, on average, 3 months [34]. This was used to estimate the progression rate from the exposed compartment to the acute infection compartment (*σ*) by estimating the average duration in the exposed compartment: 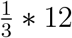. This gives a value of 4.
ix. The persistent rate of any anal HR-HPV infection among MSM was 20.6 (14.6-28.1)/1000 person-month [11]. This was used to estimate the progression rate from acute infection to persistent infection (*ρ*) by 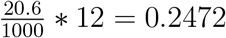 .
x. 5-year overall survival in patients who had a clinical response is 84.2% [35]. Proportion of death of 15.8% was assumed from this value. This gives a cancer-mortality rate (*ϕ*) of 0.034 using the formula 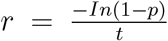, where r is the rate, p is the proportion, and t is the follow-up time.
xi. Clearance rate of any anal HR-HPV infection among MSM was 55.0 (41.0-72.3) per 1,000 person months [11]. The recovery rate from acute infection, *α*, was estimated from this value by converting it to an annual rate of 0.66.
xii. The prevalent duration of any anal HR-HPV is 23.9 months (95% CI: (21.2–26.5))[36]. This was used to estimate the recovery rate from persistent infection, *γ*, by 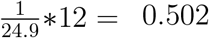.

### 4.3 Model Calibration

The least-squares method was used to optimize transmission probability (*β*_*mm*_), progression from persistent infection to HSIL (*θ*), and progression from HSIL to AC (*χ*) by calibrating these parameters to epidemiological data from the existing literature. All other model parameters are fixed at their baseline values (Table 2). *β*_*mm*_ was estimated by calibrating the model to reproduce a 40.6% reported prevalence of anal HR-HPV infection among HIV^*−*^ MSM in 2016 [14], corresponding to year 66 of the model. *θ* was estimated based on the reported HSIL prevalence of 4.4% among HIV^*−*^ MSM in 2020 [13], corresponding to year 70 of the model. *χ* was estimated using the reported anal cancer incidence rate of 19 per 100,000 person-years among HIV^*−*^ MSM in the year 2020 [15], which is year 70 of the model. This was done by minimizing the squared differences between the model outputs and the corresponding calibration targets, resulting in *β*_*mm*_ = 0.568, *θ* = 0.0161, and *χ* = 0.00432 to obtain the calibration targets simultaneously.

## 5 Results

### 5.1 Transmission Dynamics of HPV Infection Among MSM

Numerical simulation and analysis were performed using Python programming language in Google Colab to understand the transmission dynamics of HPV infection among MSM over a time interval [0, 100]. The Runge-Kutta method was used to simulate the system’s dynamics. The model was simulated with the parameters in Table 2. The initial states were set as *S*_*n*_ = 134654, *S*_*s*_ = 0, E = 1, *I*_*a*_ = 0, *I*_*p*_ = 0, *P*_*c*_ = 0, *A*_*C*_ = 0, and *R*_*HPV*_ = 0. The transmission dynamics across the compartments are shown in Figure 3.

**Figure 3:**
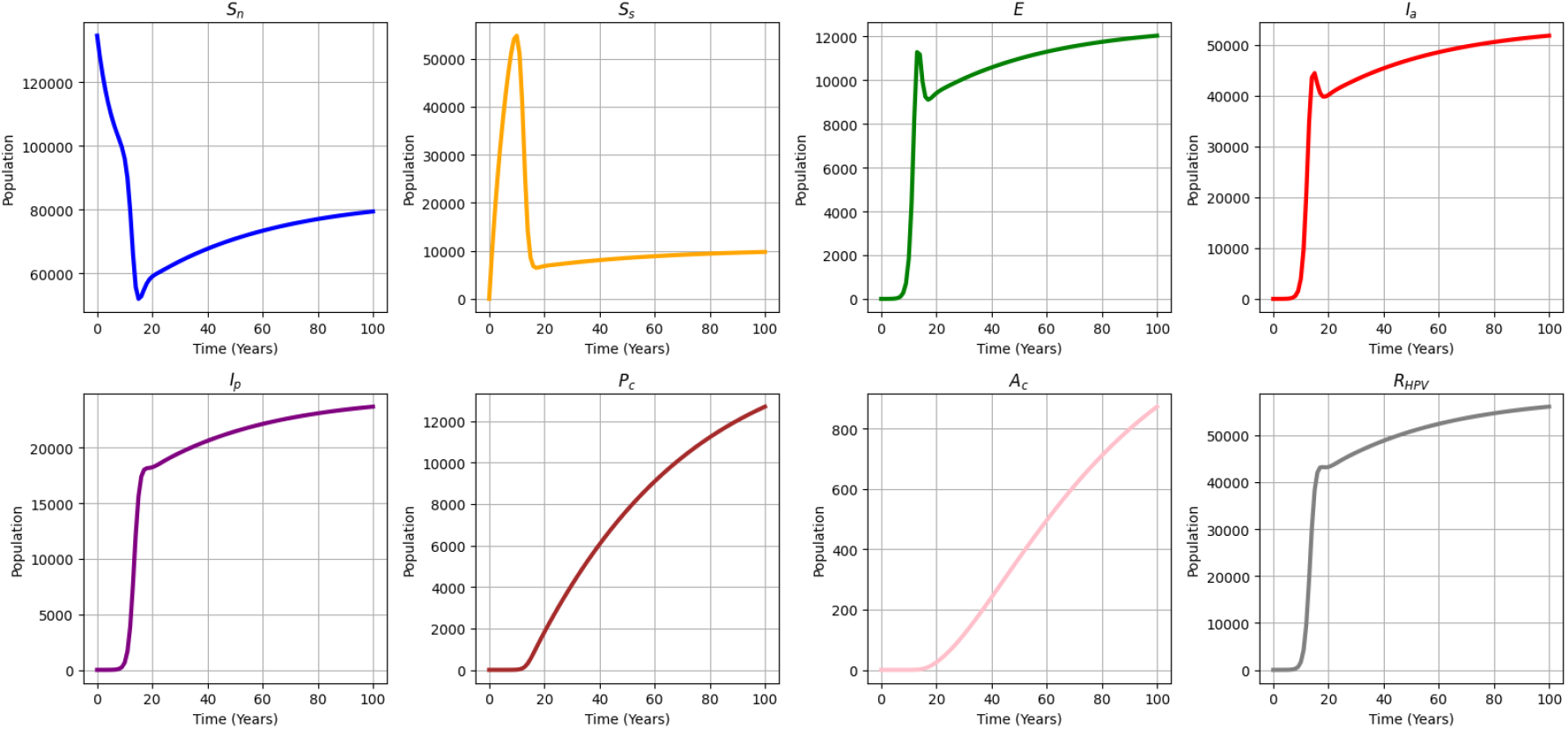
This plot illustrates the unmitigated simulation trajectories of HPV infection and disease progression over a 100-year horizon. A notable outcome is the rapid depletion of the susceptible smoker population (*S*_*s*_) within the first two decades, concurrent with an exponential increase in acute infections (*I*_*a*_), reflecting this subpopulation’s high susceptibility.

i. Susceptible Non-smokers (*S*_*n*_): The *S*_*n*_ population started with a huge population because all recruitment goes to this compartment. However, it decreased over time as individuals moved into either the Susceptible smokers (*S*_*s*_) or the Exposed (E) compartments.
ii. Susceptible Smokers (*S*_*s*_): The *S*_*s*_ compartment started with a very low population, but spikes up as transition occurs from the *S*_*n*_ compartment into it. The population crashes as the number of acute infections increases. The spike and crash in this compartment occur in the first two decades, showing how quickly the population moves into the infected compartment.
iii. Exposed (E): The exposed population starts small, peaks at around the same time as acute infection, maintains equilibrium for a while, and gradually decreases over time. The population in the exposed compartment is low compared to that in the acute compartment because the progression rate from the exposed to the acute compartment is high.
iv. Acute Infection (*I*_*a*_): The population increases from the early second decade proportional to the decline of the *S*_*s*_ and *S*_*n*_ classes. The *S*_*s*_ compartment declines less rapidly than the *S*_*n*_ compartment, and the *I*_*a*_ compartment increases rapidly, peaks in the early second decade, and continues to rise over time.
v. Persistent Infection (*I*_*p*_): The *I*_*p*_ compartment also increases as the population from the *I*_*a*_ progresses to *I*_*p*_. The peak occurs towards the late second decade and continues to rise steadily over time.
vi. HSIL (*P*_*c*_): The population with High-Grade Squamous Intraepithelial Lesion keeps a steady increase over time from the early third decade.
vii. Anal Cancer (*A*_*C*_): The progression from HSIL to *A*_*C*_ is relatively low, indicating the reason for the low population in this compartment.
viii. Recovery from HPV Infection (*R*_*HPV*_): The compartment stabilizes with the acute and persistent infection since these two compartments feed the flow into it. It shows a slight steady increase over time because the duration of infection is 1-2 years. However, the population in the compartment is lower than the *S*_*n*_ compartment because it flows into it at a faster rate, since the infection does not confer immunity among the population.

#### 5.1.1 Transmission Dynamics of the Different Susceptible Populations

Figure 4 shows the models simulated with the same initial conditions and parameter values to observe how transmission dynamics vary across populations of non-smokers and smokers susceptible (*S*_*n*_ + *S*_*s*_), only non-smokers susceptible (*S*_*n*_), and only smokers susceptible (*S*_*s*_). The (*S*_*n*_)-only and (*S*_*s*_)-only models were simulated without the smoking transition parameters *η* (transition from non-smokers to smokers) and *ϵ* (transition from smokers to non-smokers), keeping the population in a fixed smoking status over time. The force of infection (*λ*_*mm*_) in the *S*_*n*_ + *S*_*s*_ population has a maximum value of 1.007 (IQR: 0.874 - 0.905), the *λ*_*mm*_ of the *S*_*n*_-only population has a maximum value of 0.913 (IQR: 0.834 - 0.863), and the *λ*_*mm*_ of the *S*_*s*_-only population has a maximum value of 1.272 (IQR: 1.062 - 1.102). Infection starts earlier in the *S*_*s*_-only population than in the *S*_*n*_ +*S*_*s*_ population and much later in the *S*_*n*_-only population. The comparison among the three populations shows that transmission intensity is faster and the burden of disease is higher in the smokers-only population than in the non-smokers population. The *S*_*n*_ + *S*_*s*_ population shows intermediate behavior, driven by interactions between *S*_*n*_ and *S*_*s*_ and by the coupling of smoking behavior and disease dynamics. These differences reflect the impact of reduced susceptibility among non-smokers, governed by the risk-reduction parameter (*κ*), and the dynamic transitions between smoking statuses captured by the full model. This is evidence that smoking is a substantial driver for the transmission of HPV, disease progression, and long-term disease burden among MSM, and risk reduction from a non-smoking state contributes to reducing the burden of the disease in the population.

**Figure 4:**
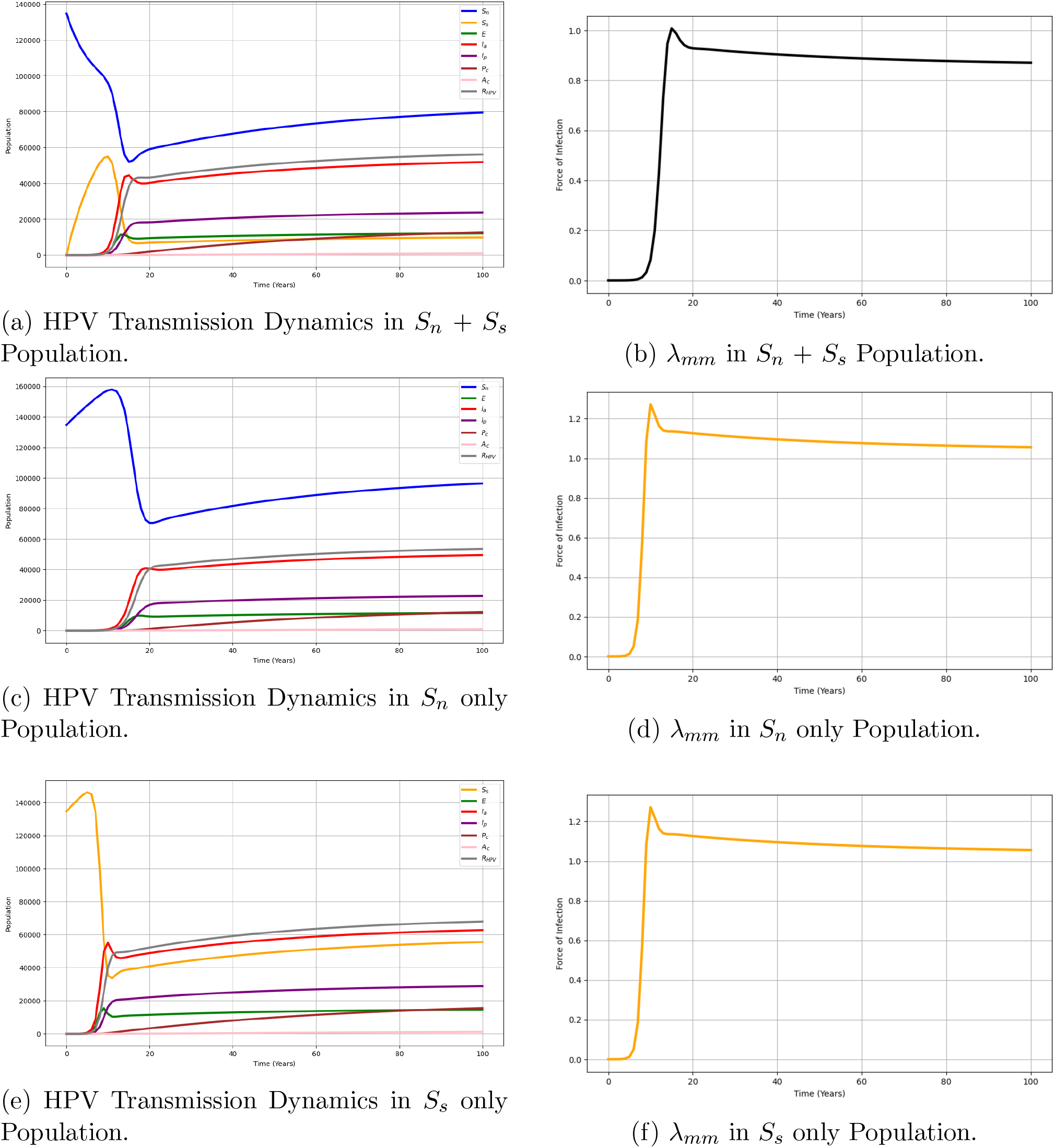
Comparison of transmission dynamics among susceptible populations. This figure contrasts the force of infection and compartmental disease burden among integrated (*S*_*n*_ + *S*_*s*_), non-smoker-only (*S*_*n*_), and smoker-only (*S*_*s*_) populations. The simulation reveals that transmission intensity peaks earlier and the long-term disease burden remains substantially higher in the smoker-only cohort compared to non-smokers, reinforcing the role of smoking as a primary catalyst for disease spread.

#### 5.1.2 Sensitivity Analysis

The model contains 16 parameters, of which 11 appear in the expression of *R*_0_. Most parameters were estimated from the existing literature, with a few estimated from the model. This is expected to lead to uncertainties in the parameter values. To understand the uncertainty and the contributions of individual and interactive behavior of the parameters, a global sensitivity analysis was conducted using Latin Hypercube Sampling-Partial Rank Correlation Coefficient (LHS-PRCC). LHS uses random sampling without replacement to analyze the entire range of each parameter by simulating N model solutions, and PRCC analyzes the strength of the relationship and the sensitivity of an outcome to variation in parameters. This sensitivity analysis identifies the parameters that significantly influence the outcome variable in the model numerical simulation [39, 40]. The range of PRCC values is -1 to 1, and higher values (*≥* 0.5) are considered a stronger correlation between the parameter and the outcome variable. A positive or negative correlation is determined by the signs of the values [41]. For this analysis, 1000 simulations per LHS were conducted, with a *±*20% range around each parameter’s baseline value, using the basic reproduction number (*R*_0_) as the outcome variable. Figure 5 shows the

**Figure 5:**
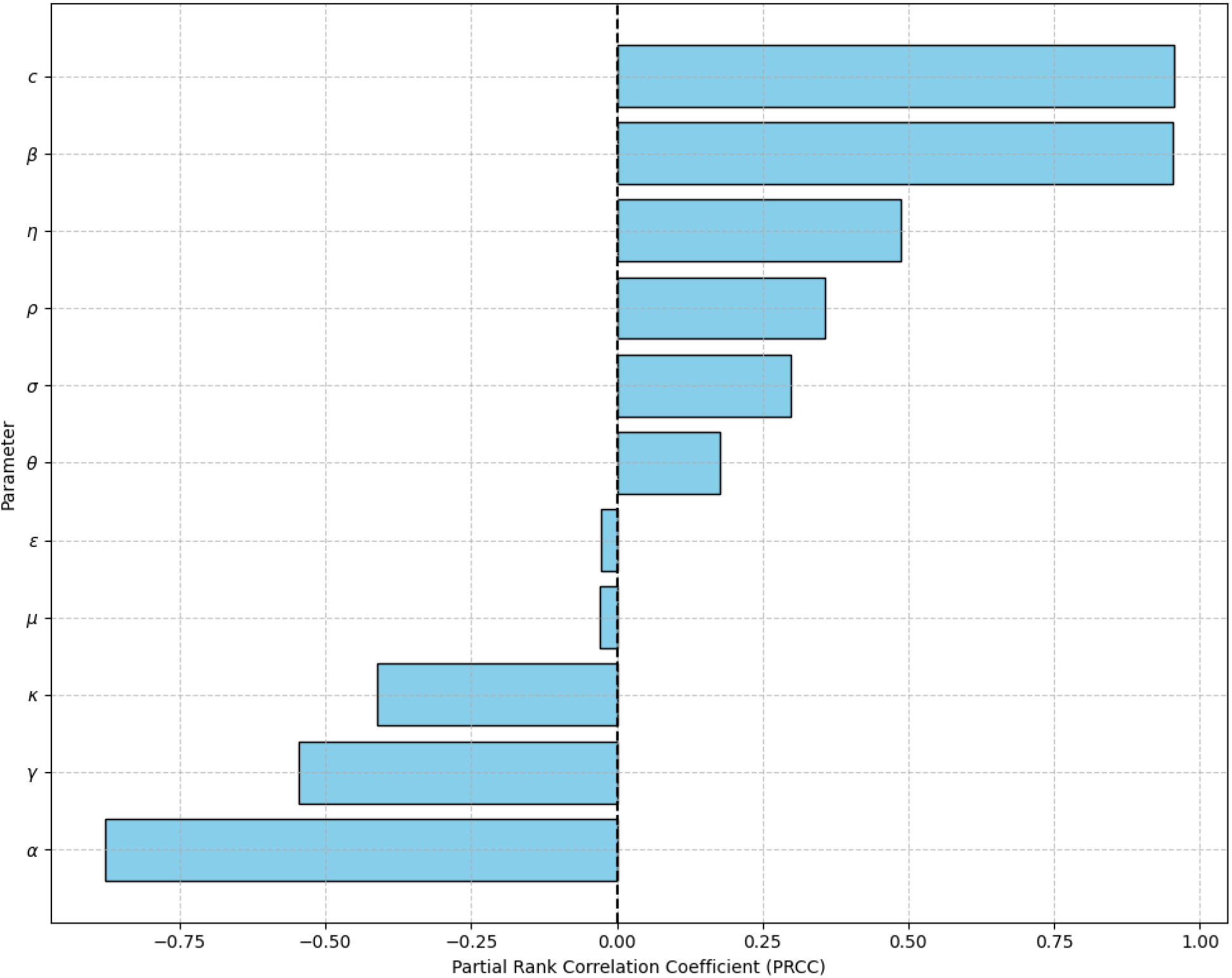
This visualization details the global sensitivity of the basic reproduction number (*R*_0_) to targeted parameter variations. The results indicate that enhancing the recovery rate from acute infection (*α*) yields the most substantial reduction in *R*_0_ (-0.879) among biological parameters, while the transition from nonsmoker to smoker (*η*) serves as a highly impactful behavioural determinant (0.487).

PRCC values indicating the direction of influence and significance of the parameters on *R*_0_, obtained from the sensitivity analysis. These parameters can be targeted to design intervention strategies to reduce the transmission and burden of HPV and AC among MSM by increasing or decreasing their values based on the direction of the relationship with the *R*_0_.

i. The average number of male contacts *c*_*mm*_ (PRCC = 0.955, p <0.001) and the transmission probability *β*_*mm*_ (PRCC = 0.953, p <0.001) have the strongest influence and are also highly significant and positively correlated with the *R*_0_. Reducing the number of male sexual contacts, engaging in safe sex practices, and raising awareness of the high risk of anal HPV transmission among MSM is pertinent in reducing the burden of HPV among MSM.
ii. The recovery from acute infection *α* (PRCC = *−*0.879, p <0.001) and recovery from persistent infection *γ* (PRCC = *−*0.546, p <0.001) also show a strong influence with a significant negative correlation with the *R*_0_. Increasing the rate of recovery from both acute and persistent infection through screening and treatment of infected MSM will reduce the transmission of the disease. Screening services should be made available, and MSM should be encouraged to know their HPV status for early detection and prompt treatment to promote recovery and reduce the possibilities of disease progression.
iii. The transition from nonsmoker to smoker *η* (PRCC = 0.487, p <0.001) shows a moderate influence on the *R*_0_ with a significant positive correlation, while the reduced susceptibility due to nonsmoking status *κ* (PRCC = *−*0.412, p <0.001) has a significant negative correlation with moderate influence on the *R*_0_. This shows how smoking drives HPV transmission and the efficacy of the protective effect of nonsmoking on HPV acquisition and transmission. In combination with other preventive strategies, advocating for a reduction in smoking, most importantly, preventing smoking initiation and relapse among the MSM population, will help to reduce the transmission, progression, and burden of the disease in the population.
iv. The progression from acute to persistent infection *ρ* (PRCC = 0.356, p <0.001), progression from exposed to acute infection *σ* (PRCC = 0.299, p <0.001), and progression from persistent infection to HSIL *θ* (PRCC = 0.176, p <0.001) have a significant positive correlation with the *R*_0_, though with a weak influence. There is a need for proper treatment of MSM with HPV to reduce the progression of the disease and promote recovery in the population.
v. The natural death rate *µ* (PRCC = -0.029, p = 0.359) and the transition from nonsmoker to smoker *ϵ* (PRCC = -0.028, p = 0.359) both show a very weak influence and no significant negative correlation with *R*_0_.

#### 5.1.3 Sensitivity Analysis of Smoking Reduction on Transmission Dynamics of HPV Among MSM

The model examines the reduction in transition from non-smokers to smokers (*η*) and the increase in transition from smokers to non-smokers (*ϵ*) as a smoking reduction intervention to identify how it influences the transition dynamics of HPV and AC progression among MSM. The model estimates how reductions of 25%, 50%, 75%, and 100% in *η* and the corresponding increases in *ϵ* jointly impact the transmission dynamics of HPV and anal cancer burden. Figure 6 shows that all intervention levels increase the *S*_*n*_ population while reducing the populations in other compartments. This indicates that smoking reduction can help to reduce the burden of HPV and anal cancer among MSM.

**Figure 6:**
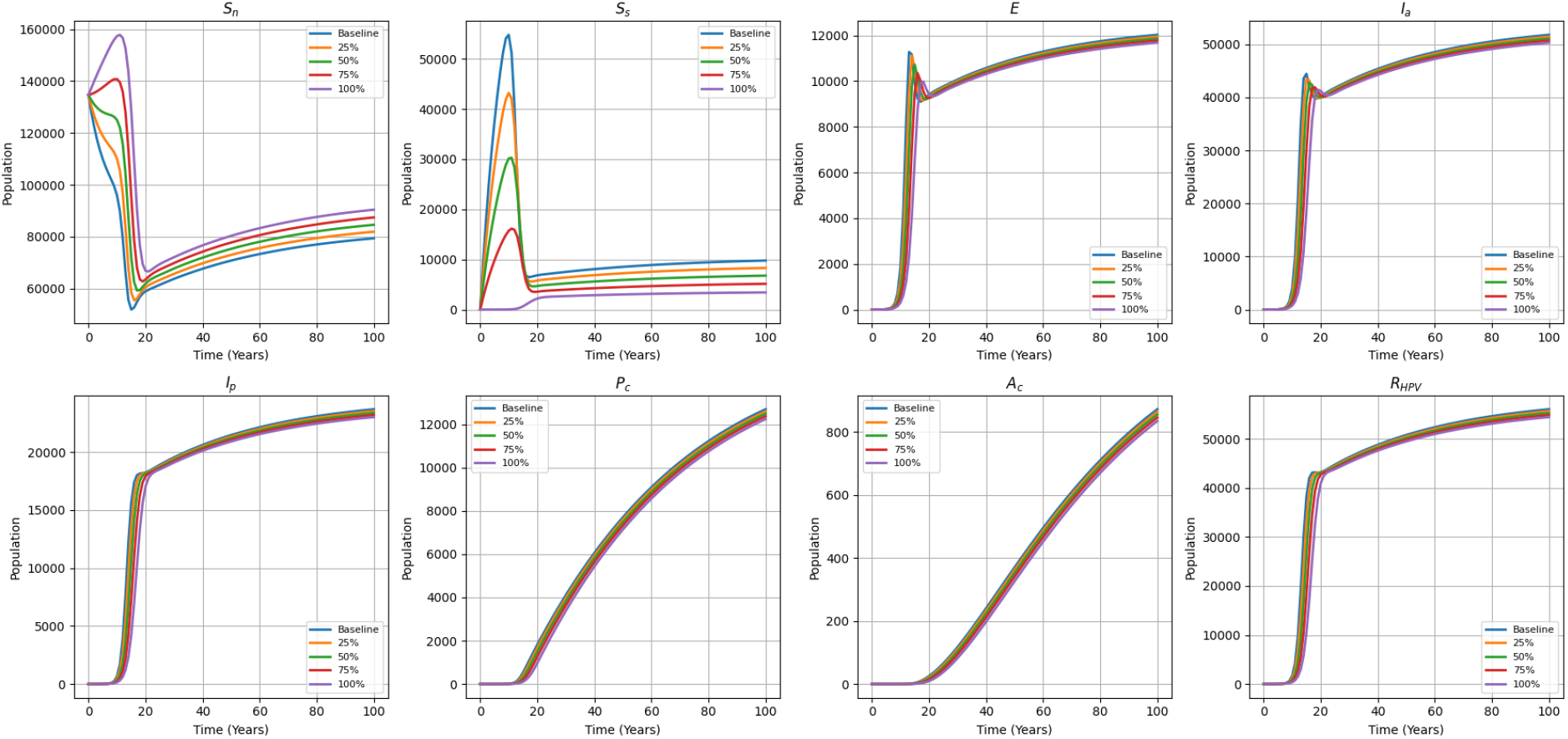
These trajectories project the epidemiological impact of varying smoking-reduction efficiencies (25% to 100%) relative to baseline disease progression. The simulations emphasize that while aggressive smoking reduction mitigates overall disease incidence and expands the non-smoker pool, this intervention alone is insufficient to eradicate the disease burden.

### 5.2 Interventions

#### 5.2.1 Impacts of Smoking Reduction on Basic Reproduction Number

The parameters given in Table 2 were used to compute the basic reproduction number (*R*_0_) in the MSM population comprising non-smokers and smokers, yielding a value of 3.54 (95% UI: 2.41 - 5.01). This shows that without intervention, HPV transmission and anal cancer will persist in the population. However, reducing smoking by 25%, 50%, 75%, and 100% reduces *R*_0_ by 5.93%, 12.42%, 19.67%, and 28.48%, respectively, with *R*_0_ values of 3.33 (95% UI: 2.25 - 4.72), 3.09 (95% UI: 2.08 - 4.39), 2.84 (95% UI: 1.90 - 4.05), and 2.53 (95% UI: 1.67 - 3.69), respectively. With this, there will be a reduction in the burden of the disease, although the disease will persist since 100% smoking reduction does not reduce *R*_0_ below the threshold, indicating its insufficiency to reduce HPV transmission and burden alone. The individual effects of reducing the transition from non-smoker to smoker, increasing the transition from smoker to non-smoker, and the joint effect of the two at 25%, 50%, 75%, and 100% were compared to see the influence on *R*_0_. The synergistic effect of the two scenarios has a greater impact on *R*_0_, underscoring the need for preventive strategies targeting smoking initiation and cessation to reduce disease transmission and burden. Reducing the transition from non-smoker to smoker has more influence on *R*_0_ compared to increasing the transition from smoker to non-smoker. This shows how preventing smoking initiation and relapse after cessation can be substantial in the preventive intervention strategies.

#### 5.2.2 Joint Analysis of Intervention with Vaccination Coverage and Contact Reduction

To further explore control strategies aimed at disease control, the critical vaccination coverage required to reach the traditional Herd Immunity Threshold (HIT) was estimated using the basic reproduction number (*R*_0_) and the vaccine efficacy (*E*_*v*_). The Gardasil vaccine is used in Nigeria for its HPV immunization program [42] with an efficacy of 75% against Anal Cancer in Males [43]. The Herd Immunity Threshold is defined as the proportion of the susceptible population that falls below the level required for disease transmission [44]. The HIT is derived by using the relationship of Herd Immunity Threshold (HIT) to *R*_0_, where:

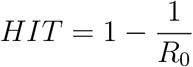

Since the vaccine does not guarantee perfect protection, the HIT was subsequently adjusted for 75% vaccine efficacy (*E*_*v*_) to achieve the required vaccine coverage *V*_*c*_ to interrupt HPV transmission. The vaccine reproduction number (*R*_*v*_) was set to 1 to solve for the critical vaccination coverage needed to achieve HIT (0 < *E*_*V*_ < 1 and 0 *≤V*_*C*_ *≤* 1). Therefore:

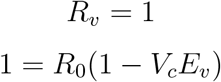

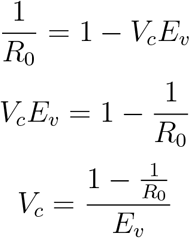

The estimates of the synergistic impact of vaccination and smoking reduction indicate that the greater the percentage of smoking reduction, the lower the required vaccine coverage. In the absence of smoking reduction, 93.3% (76.8–99.6) vaccination coverage is required, compared with 80.6% (53.7–96.8) at 100% smoking reduction. Analysis of the synergistic effect of smoking and contact reduction shows that, in the absence of smoking reduction, approximately 71.8% (58.5–80.1) contact reduction is required to reduce the *R*_0_ below unity, compared with 60.5% (40.4–73.0) at 100% smoking reduction (Figure 7 and Table 3). This indicates that the combined strategies reduce the level of vaccination coverage and behaviour modification essential to control the transmission and burden of the diseases. MSMs should be a priority target population for HPV vaccination. While reaching HIT brings *R*_0_ below unity, the presence of backward bifurcation implies that this coverage must be maintained or exceeded to overcome endemic persistence and inform subsequent vaccination interventions among the MSM population.

**Table 3:**
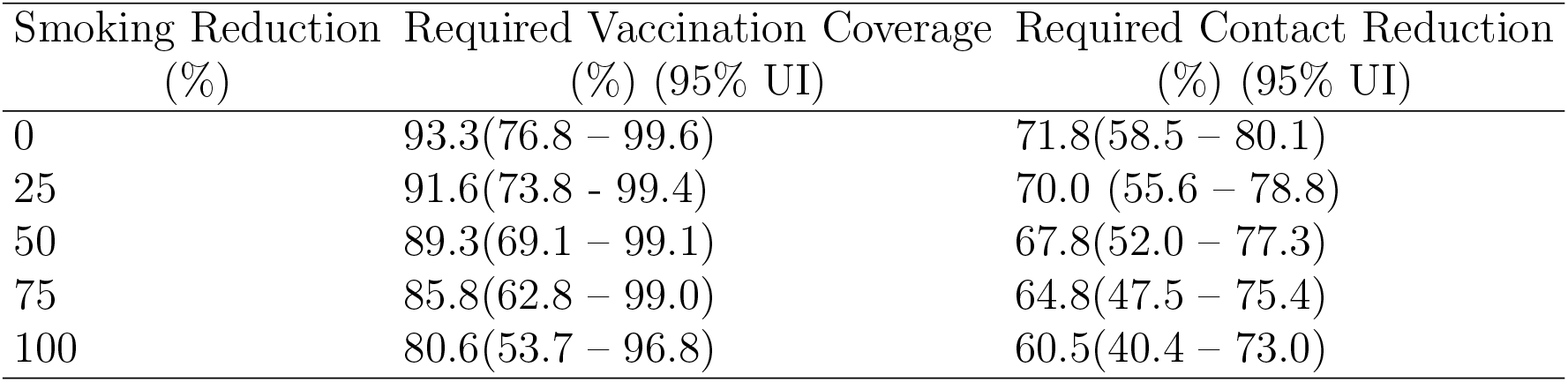
The table shows the required critical vaccination coverage and contact reduction necessary to drive *R*_0_ below unity. As indicated by the model’s backward bifurcation, reaching this sub-unity threshold is a necessary prerequisite, though it may require synergistic interventions of even greater magnitude to achieve complete elimination if the initial disease burden is high.

| Smoking Reduction<br>(%) | Required Vaccination Coverage<br>(%) (95% UI) | Required Contact Reduction<br>(%) (95% UI) |
| --- | --- | --- |
| 0 | 93.3(76.8 – 99.6) | 71.8(58.5 – 80.1) |
| 25 | 91.6(73.8 – 99.4) | 70.0 (55.6 – 78.8) |
| 50 | 89.3(69.1 – 99.1) | 67.8(52.0 – 77.3) |
| 75 | 85.8(62.8 – 99.0) | 64.8(47.5 – 75.4) |
| 100 | 80.6(53.7 – 96.8) | 60.5(40.4 – 73.0) |

**Figure 7:**
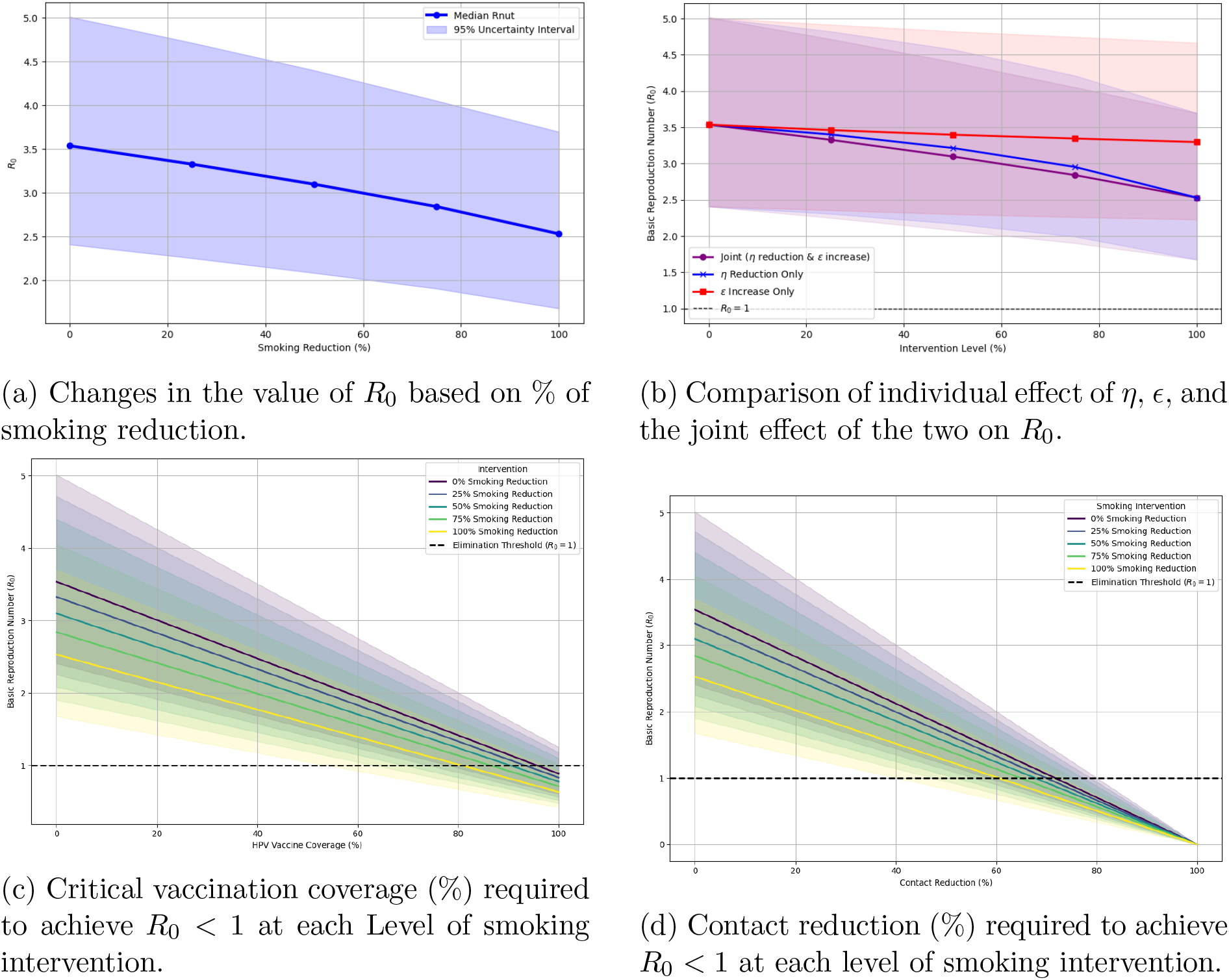
This figure depicts the reduction in *R*_0_ under progressive smoking cessation policies, cross-evaluated with required vaccination coverage and reductions in sexual contact rates. Crucially, the analysis demonstrates that a 100% smoking reduction only suppresses *R*_0_ to 2.53; reducing the epidemic threshold below unity necessitates a concurrent 80.6% critical vaccination coverage or 60.5% decrease in contact rates, underscoring the requirement for combination prevention strategies.

#### 5.2.3 Impact of Smoking reduction on Incidence and Prevalence (2026 – 2046)

Model estimates show how variation in smoking-reduction rates modifies incidence and prevalence of HPV and AC trends relative to the baseline. Although the incidence and prevalence continue to increase over the years, the intervention reduces the expected burden over the study period. This shows the need for early implementation of interventions before there is a substantial accumulation of HPV-infected, precancerous, and cancer cases. A 100% smoking-reduction intervention reduced the projected HPV prevalence by 2.60% compared with the projected 0.58% increase from 2026 to 2046; there is also a 3.16% reduction relative to the 2046 baseline projection. HPV incidence was reduced by 1% compared to a 2% increase between the years, and by 3% relative to the 2046 baseline value. The projected increase in AC prevalence between the years was 20.91% and was reduced to 15.15%; also, there is a 4.76% reduction in the 2046 baseline projection. The projected AC incidence estimate was reduced from 13.04% to 9% between the years, and a 5% reduction relative to the 2046 baseline projection (Table 4).

**Table 4:** This table presents the epidemiological efficacy of varying smoking reduction interventions over 20 years (2026–2046). The projections establish that optimal smoking reduction interventions consistently avert a significant volume of new infections compared to the unmitigated baseline, highlighting the importance of behavioral modification in controlling HPV burden.

| Metric | Baseline Year 2026<br>(Median, 95% UI) | Baseline Year 2046<br>(Median, 95% UI) | 100% Intervention Year 2046<br>(Median, 95% UI) |
| --- | --- | --- | --- |
| HPV Prevalence | 0.4039<br>(0.2900, 0.4954) | 0.4063<br>(0.2927, 0.4976) | 0.3934<br>(0.2602, 0.4925) |
| HPV Incidence | 46087<br>(34924, 57734) | 47082<br>(35557, 59862) | 45849<br>(32052, 59051) |
| AC Prevalence | 0.0028<br>(0.0016, 0.0043) | 0.0034<br>(0.0020, 0.0051) | 0.0032<br>(0.0017, 0.0050) |
| AC Incidence | 46<br>(27, 73) | 52<br>(31, 85) | 50<br>(27, 82) |

The comparison plots of the different susceptible populations show that the prevalence and incidence of HPV and AC are higher over time in the smokers-only population compared to the other populations. These findings highlight that the intervention reduces the burden of the disease over time, indicating the role that smoking plays in sustaining susceptibility, transmission, and progression of the disease (Figure 8).

**Figure 8:**
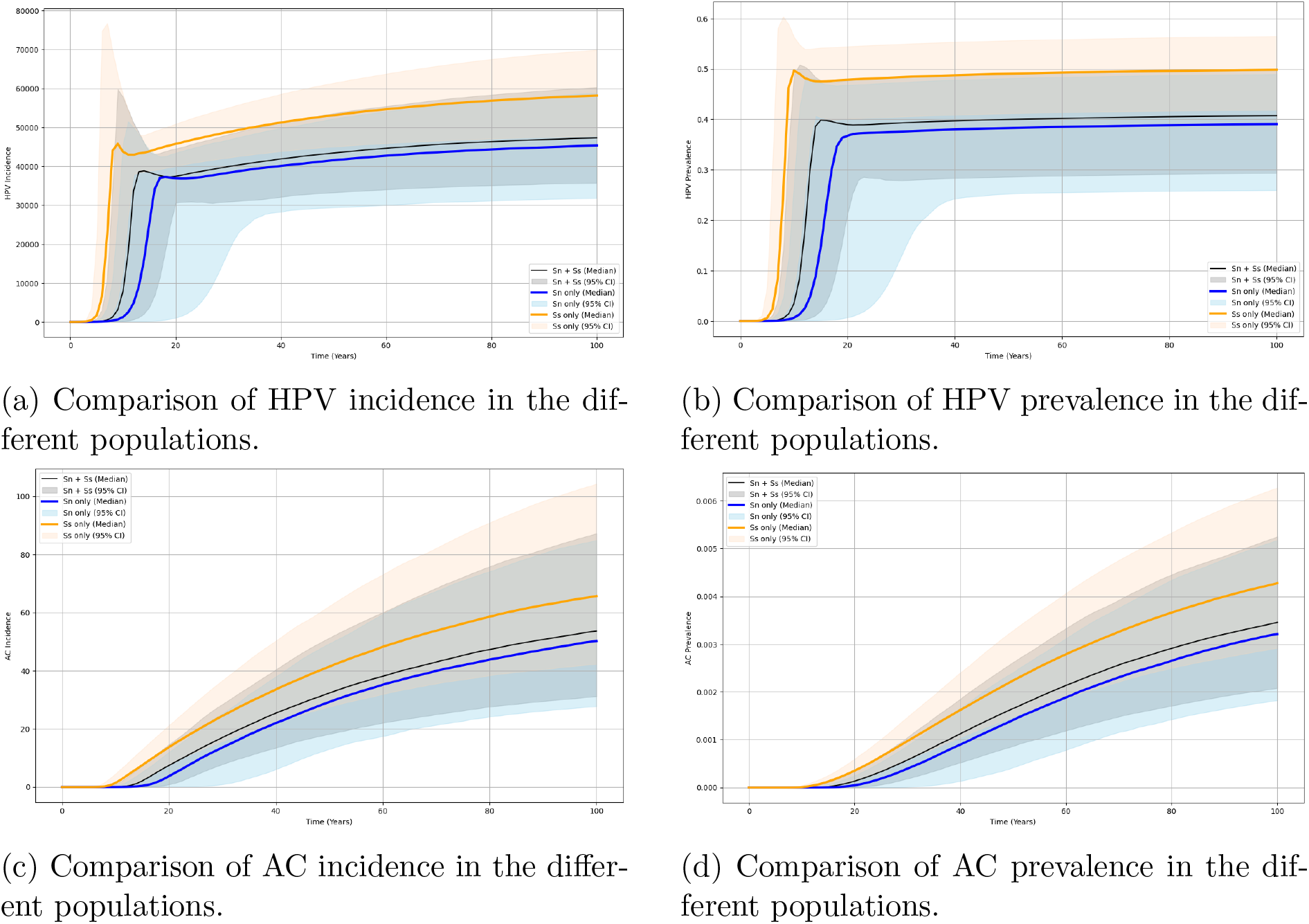
These plots show the differences in incidence and prevalence of HPV and AC in the different populations. Incidence and Prevalence of HPV and AC are higher in the *S*_*s*_ only population, highlighting the role of smoking in increasing susceptibility, transmission, and progression of the disease.

#### 5.2.4 Comparison of HPV Incidence and AC Incidence and Prevalence Based on HIV Status

The HIV^+^ MSM population is at a higher risk of HPV infection and AC compared to the HIV^*−*^ MSM population. The baseline parameters (Table 2) were modified to compare the impact of HIV-HPV co-infection and how the prevalence of HPV, AC incidence, and AC prevalence differ from the HIV^*−*^ MSM population. HIV^+^ MSM had a higher persistence (16.7% vs. 1.3%, p<0.001) of HPV 16 than HIV^*−*^ MSM [45]. This was used to estimate the persistence rate of any anal HR-HPV (*ρ*_*HIV*_) among HIV^+^ MSM (*ρ*_*HIV*_ = (16.7/1.3) ^*^ baseline *ρ*). The progression rate from persistent infection to HSIL (*θ*) was multiplied by 1.77 to obtain *θ*_*HIV*_, corresponding to the ratio of HSIL prevalence in HIV^+^ MSM (7.8%) to that in HIV^*−*^ (4.4%) [13]. The incidence rate of anal cancer in HIV^+^ MSM is 89/100,000 person-years compared to 19/100,000 person-years in HIV^*−*^ MSM [15]. The progression rate from HSIL to anal cancer for the HIV^+^ MSM (*χ*_*HIV*_) was estimated by multiplying the baseline *χ* by 4.47. The outcomes show that HPV prevalence and AC incidence and prevalence are noticeably higher in the HIV^+^ MSM population compared to the HIV^*−*^ MSM population (Figure 9).

**Figure 9:**
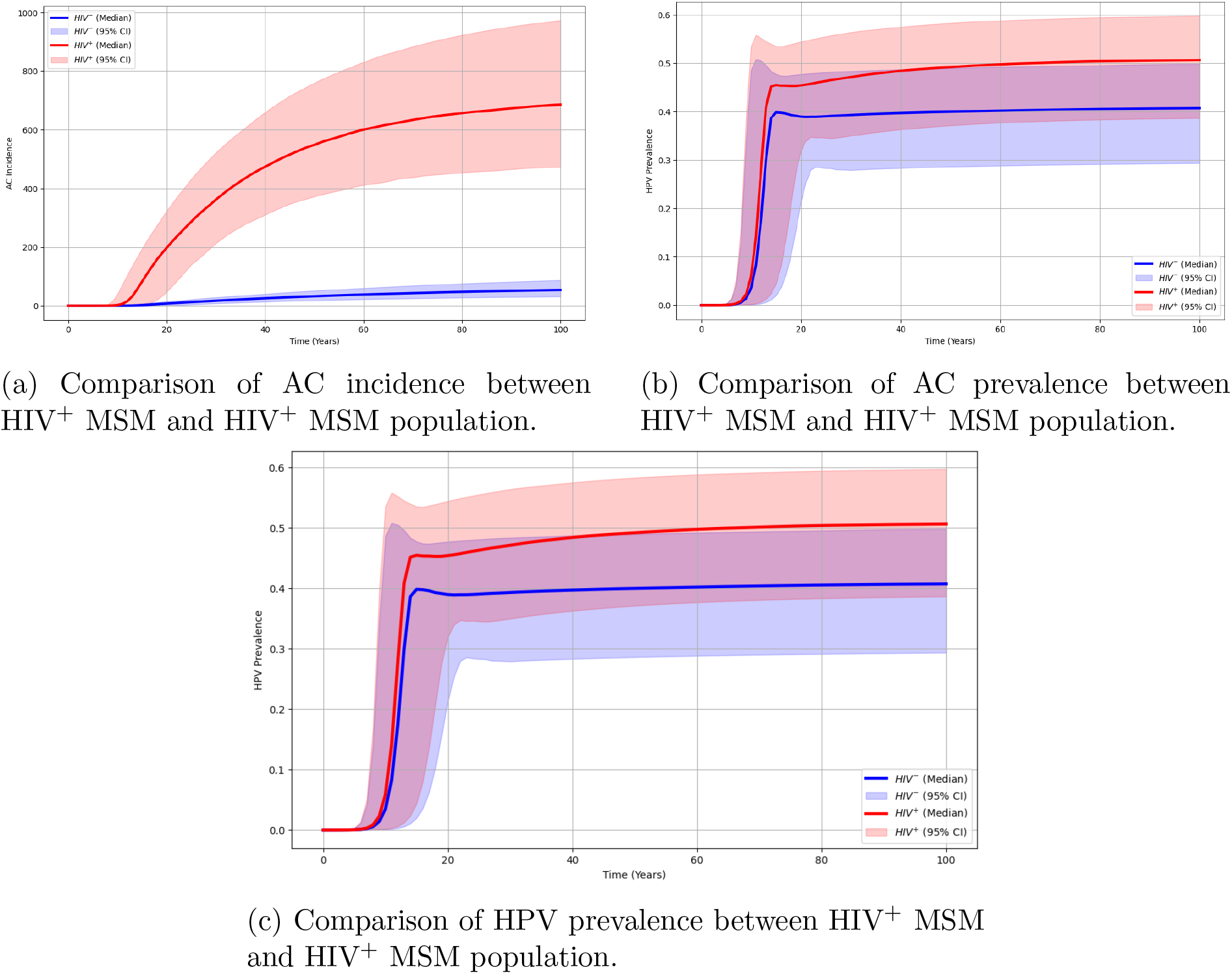
The comparison between AC incidence and AC and HPV prevalence among the MSM based on HIV status shows that the disease burden is higher among the HIV^+^ MSM than that in the HIV^*−*^ MSM. This indicates an increased susceptibility to HPV infection and the need for the HIV^+^ MSM to be a priority target for public health interventions.

## 6 Discussion

This study shows how smoking reduction influences the transmission dynamics of HPV among MSM. The analysis yields biologically and epidemiologically meaningful inferences, as evidenced by the impact of smoking reduction on the force of infection, *R*_0_, the incidence and prevalence of the disease. The LHS-PRCC sensitivity analysis of the model parameters with respect to *R*_0_ shows that the contact rate, transmission probability, transition from non-smoker to smoker, and disease progression parameters have a significant positive correlation with *R*_0_, while the recovery parameters and the protective effect of non-smoking status have a significant negative correlation with *R*_0_. The analysis of the force of infection shows a higher value in the smoker-only population compared to the non-smoker population. This is consistent with the finding that current smokers have a higher susceptibility to genital HPV infection compared to non-smokers [46, 17]. Smoking reduction alone cannot reduce *R*_0_ below the threshold necessary to curb the spread of the disease; however, the joint analysis of smoking reduction and vaccination, and of smoking reduction and contact reduction, brought *R*_0_ < 1, demonstrating how the interplay between interventions reduces the level of vaccination coverage and behavioral change required for disease prevention and control. Nevertheless, backward bifurcation in the model complicates such a strategy. Since an endemic equilibrium and a disease-free equilibrium can coexist even when *R*_0_ < 1, reducing *R*_0_ slightly below 1 is insufficient to eliminate the infection from the population in the presence of a large disease burden. In this case, there is an urgent need to adopt early and effective combination strategies to reduce the reproduction number far below the bifurcation point and eradicate HPV among MSM. Smoking plays a pivotal role in overall transmission dynamics by increasing susceptibility and sustaining higher infection levels in the population; contact drives transmission intensity; and vaccination is required to protect the entire susceptible population from HPV infection. Therefore, integrated behavioral interventions and vaccination will have a synergistic effect on reducing HPV transmission among MSM. Behavioral interventions such as smoking cessation should be integrated into anal cancer prevention programs to reduce the risk of HR-HPV persistence [45].

Smoking increases the incidence and prevalence of HPV and anal cancer due to increased susceptibility to the disease; however, the burden is reduced over time by the interventions compared to baseline values without intervention. The comparison plots of susceptible populations show that the burden of HPV and anal cancer is highest in the smoker-only population. This finding is supported by the report that smoking reduction is likely to increase the resolution of HPV infection compared to frequent, long-term, or heavy smoking [18]. The synergistic impact of the smoking parameters on *R*_0_ is stronger than the impact of each parameter individually, emphasizing the need to discourage smoking initiation and relapse, and to encourage smoking cessation in the population. The transition from non-smokers to smokers has a greater influence on disease transmission and burden than the transition from smokers to non-smokers. The HIV^+^ MSM population is projected to have a higher incidence of HPV and AC, and a higher AC prevalence, compared with the HIV^*−*^ MSM population, consistent with the finding that HIV^+^ MSM have a higher prevalence of anal HR-HPV, multiple anal HR-HPV types, and HSIL compared to HIV^*−*^ MSM [14, 13]. The incidence rate of anal cancer among HIV-positive MSM is substantially higher than among HIV-negative MSM (85 vs. 19 per 100,000 person-years) [15], and HIV^+^ MSM are at higher risk of anal high-risk HPV persistence [45]. The HIV^+^ MSM population is therefore at higher risk of disease progression and burden, and should be a priority target for public health interventions. There is a need for integrated interventions to curtail the transmission and burden of disease in high-risk populations, particularly in countries where vaccination is unavailable for the male population. The findings of this study will contribute to the development of targeted interventions among MSM to reduce the incidence and burden of disease in both specific and general populations.

However, this modelling study has several limitations. A major limitation is the lack of epidemiological data on anal HR-HPV among MSM in Nigeria, which necessitated the use of regional or global data from the literature to estimate model parameters. The optimized parameters *β*_*mm*_, *θ*, and *χ* were each calibrated using a single data point. The model adopts a homogeneous mixing assumption, whereas MSM populations exhibit well-documented assortative mixing patterns (e.g., by age, HIV status, and risk behavior). In addition, the MSM population is modeled as closed and male-only, yet HPV transmission can involve bisexual men who bridge to female populations. Smoking and non-smoking are treated as persistent statuses across all compartments that modify HPV and cancer risk, and the model considers only the effect of smoking behavior on susceptibility to HPV infection.

## 7 Conclusion

MSM require targeted interventions due to their high susceptibility to HPV infection. Behavioral interventions such as smoking reduction will contribute to reductions in the incidence and prevalence of HPV and AC. A 100% smoking-reduction intervention reduced the projected HPV prevalence by 2.60% compared with a projected increase of 0.58% from 2026 to 2046, and by 3.16% relative to the 2046 baseline projection. HPV incidence was reduced by 1% compared to a projected increase of 2% over the same period, and by 3% relative to the 2046 baseline value. The projected increase in AC prevalence over the period was 20.91%, which was reduced to 15.15%; there was also a 4.76% reduction relative to the 2046 baseline projection. The projected AC incidence was reduced from 13.04% to 9% over the period, with a 5% reduction relative to the 2046 baseline projection. Furthermore, with 100% smoking reduction, the basic reproduction number *R*_0_ decreased from 3.54 to 2.53, indicating reduced disease transmission. Combining smoking reduction with vaccination reduces the critical vaccination coverage required to drive *R*_0_ below unity from 92.8% (78.4-99.3%) to 80.4% (55.7-98.4%) at 100% smoking reduction. Furthermore, the synergistic effect of smoking and contact reduction lowers the contact reduction required to cross the traditional elimination threshold (*R*_0_ < 1) from 71.8% (58.5-80.1%) to 60.5% (40.4-73.0%). However, due to backward bifurcation, policymakers must treat these targets as minimum requirements, necessitating sustained behavioral changes to completely prevent and control the disease among MSM. Concurrent interventions targeting susceptible populations will sufficiently reduce HPV transmission, thereby lowering the incidence, long-term burden, and complications of HPV and HPV-related cancers in the population. It is recommended that health authorities intensify efforts to reduce smoking by raising awareness of its harms to individual health and broader public well-being.

## Data Availability

All data produced in the present work are contained in the manuscript.

## LIST OF ABBREVIATIONS

AC: Anal Cancer
aOR: Adjusted Odds Ratio
CI: Confidence Interval
HGAIN: High Grade Anal Intraepithelial Neoplasia
HPV: Human Papillomavirus
Hr-HPV: High Risk Human Papillomavirus
HSIL: High Grade Squamous Intraepithelial Lesion
LMICs: Low-and Middle-Income Countries
MSM: Men who have Sex with Men
NGM: Next Generation Matrix
ODE: Ordinary Differential Equations
UI: Uncertainty Interval
R_0_: Basic Reproduction Number

## Acknowledgements

ROO is supported by PhD fellowship from Ahmedabad University.

## Author Contributions

Rashidat Oluwabukola Owolabi: Conceptualization, Methodology, Software, Data curation, Visualization, Writing-Original draft. Maia Martcheva: Methodology, Formal analysis, Supervision, Writing-Original draft. Indrajit Ghosh: Conceptualization, Methodology, Supervision, Project administration, Writing-Original draft.

## Declaration of generative AI and AI-assisted technologies in the manuscript preparation process

During the preparation of this work, the author(s) used Claude Sonnet to typeset handwritten analysis into a LaTeX file. After using this tool, the author(s) reviewed and edited the content as needed and take(s) full responsibility for the content of the published article.

## References

[1] Karthick Nithyanandhan, Shoba Mammen, and Priya Abraham. “Human papilomavirus infection in non-cervical sites in India”. In: Indian Journal of Medical Microbiology 57 (Sept. 1, 2025), p. 100948. DOI: 10.1016/j.ijmmb.2025.100948.

[2] Anna R. Giuliano, Gabriella Anic, and Alan G. Nyitray. “Epidemiology and pathology of HPV disease in males”. In: Gynecologic oncology 117.2 0 (May 2010), S15–S19. DOI: 10.1016/j.ygyno.2010.01.026.

[3] Kathrine D. Lycke et al. “An updated understanding of the natural history of cervical human papillomavirus infection—clinical implications”. In: American Journal of Obstetrics and Gynecology 232.5 (May 1, 2025), pp. 453–460. DOI: 10.1016/j.ajog.2025.02.029.

[4] HPV and Cancer - NCI. Mar. 1, 2019. URL: https://www.cancer.gov/about-cancer/causes-prevention/risk/infectious-agents/hpv-and-cancer (vis-ited on 04/10/2026).

[5] Soumendu Patra et al. “HPV and Male Cancer: Pathogenesis, Prevention and Impact”. In: Journal of the Oman Medical Association 2.1 (June 2025), p. 4. DOI: 10.3390/joma2010004.

[6] CDC. Chapter 11: Human Papillomavirus. Epidemiology and Prevention of Vaccine-Preventable Diseases. Jan. 27, 2026. URL: https://www.cdc.gov/pinkbook/hcp/table-of-contents/chapter-11-human-papillomavirus.html (visited on 03/30/2026).

[7] Laia Bruni et al. “Global and regional estimates of genital human papillomavirus prevalence among men: a systematic review and meta-analysis”. In: The Lancet Global Health 11.9 (Sept. 1, 2023), e1345–e1362. DOI: 10.1016/S2214-109X(23)00305-4.

[8] Natália Luiza Kops et al. “Prevalence and correlates of anal HPV infection among men who have sex with men in Brazil: A respondent-driven sampling study”. In: International Journal of Infectious Diseases 163 (Feb. 1, 2026), p. 108292. DOI: 10.1016/j.ijid.2025.108292.

[9] E. S. L. Pedersen, D. Verschoor, and E. Segelov. “Incidence and burden of anal cancer— time to fight the growing disparities”. In: ESMO Gastrointestinal Oncology (Feb. 24, 2025), p. 100147. DOI: 10.1016/j.esmogo.2025.100147.

[10] Zewen Zhang et al. “Natural History of Anal Papillomavirus Infection in HIV-Negative Men Who Have Sex With Men Based on a Markov Model: A 5-Year Prospective Cohort Study”. In: Frontiers in Public Health 10 (May 11, 2022), p. 891991. DOI: 10.3389/fpubh.2022.891991.

[11] Xinyi Zhou et al. “Incidence, persistence, and clearance of anogenital human papillomavirus among men who have sex with men in Taiwan: a community cohort study”. In: Frontiers in Immunology 14 (June 20, 2023), p. 1190007. DOI: 10.3389/fimmu.2023.1190007.

[12] Sofie H. Mooij et al. “No evidence for a protective effect of naturally induced HPV antibodies on subsequent anogenital HPV infection in HIV-negative and HIV-infected MSM”. In: Journal of Infection 69.4 (Oct. 1, 2014), pp. 375–386. DOI: 10.1016/j.jinf.2014.06.003.

[13] Rebecca G. Nowak et al. “Multiple HPV infections among men who have sex with men engaged in anal cancer screening in Abuja, Nigeria”. In: Papillomavirus Research 10 (Dec. 1, 2020), p. 100200. DOI: 10.1016/j.pvr.2020.100200.

[14] Rebecca G. Nowak et al. “Prevalence of Anal High-Risk Human Papillomavirus Infections Among HIV-Positive and HIV-Negative Men Who Have Sex With Men in Nigeria”. In: Sexually Transmitted Diseases 43.4 (Apr. 2016), p. 243. DOI: 10.1097/OLQ.0000000000000431.

[15] Gary M. Clifford et al. “A meta-analysis of anal cancer incidence by risk group: Toward a unified anal cancer risk scale”. In: International Journal of Cancer 148.1 (2021), pp. 38–47. DOI: 10.1002/ijc.33185.

[16] Glenn-Milo Santos et al. “Demographic, Behavioral, and Social Characteristics Associated With Smoking and Vaping Among Men Who Have Sex With Men in San Francisco”. In: American Journal of Men’s Health 13.3 (May 1, 2019), p. 1557988319847833. DOI: 10.1177/1557988319847833.

[17] Victoria Umutoni et al. “The Association Between Smoking and Anal Human Papillomavirus in the HPV Infection in Men Study”. In: Cancer epidemiology, biomarkers & prevention : a publication of the American Association for Cancer Research, cosponsored by the American Society of Preventive Oncology 31.8 (Aug. 2, 2022), pp. 1546–1553. DOI: 10.1158/1055-9965.EPI-21-1373.

[18] Kangli Ma et al. “Impact of smoking exposure on human papillomavirus clearance among Chinese women: A follow-up propensity score matching study”. In: Tobacco Induced Diseases 21 (Mar. 20, 2023), p. 42. DOI: 10.18332/tid/161026.

[19] Soyoung Park, Hyunah Lim, and Abba B. Gumel. “Mathematical Assessment of the Roles of Vaccination and Pap Screening on the Burden of HPV and Related Cancers in Korea”. In: Bulletin of Mathematical Biology 87.12 (Dec. 3, 2025), p. 182. DOI: 10.1007/s11538-025-01548-5.

[20] Henok Desalegn Desta et al. “Mathematical Model of Human Papillomavirus (HPV) Dynamics With Double-Dose Vaccination and Its Impact on Cervical Cancer”. In: Discrete Dynamics in Nature and Society 2024.1 (Jan. 2024). Ed. by Francisco R. Villatoro, p. 9971859. ISSN: 1026-0226, 1607-887X. DOI: 10.1155/ddns/9971859. URL: https://onlinelibrary.wiley.com/doi/10.1155/ddns/9971859 (visited on 03/25/2026).

[21] Jane J. Kim. “Mathematical Model of HPV Provides Insight into Impacts of Risk Factors and Vaccine”. In: PLOS Medicine 3.5 (Apr. 4, 2006), e164. ISSN: 1549-1676. DOI: 10.1371/journal.pmed.0030164. URL: https://journals.plos.org/plosmedicine/article?id=10.1371/journal.pmed.0030164 (visited on 05/02/2026).

[22] A. Omame, D. Okuonghae, and S. C. Inyama. “A Mathematical Study of a Model for HPV with Two High-Risk Strains”. In: Mathematical Modelling in Health, Social and Applied Sciences. Ed. by Hemen Dutta. Singapore: Springer, 2020, pp. 107–149. ISBN: 978-981-15-2286-4. DOI: 10.1007/978-981-15-2286-4_4. URL: https://doi.org/10.1007/978-981-15-2286-4_4 (visited on 05/02/2026).

[23] Ogechi Regina Amanso et al. “A novel mathematical model for transmission dynamics of HPV and cervical cancer progression with cancer-reliant awareness”. In: Journal of the Nigerian Society of Physical Sciences (Mar. 22, 2026). ISSN: 2714-4704. DOI: 10.46481/jnsps.2026.3224. URL: https://journal.nsps.org.ng/index.php/jnsps/article/view/3224 (visited on 05/02/2026).

[24] Ogechi Regina Amanso et al. “Analysis of a mathematical model of human papillomavirus transmission dynamics with optimal control for cervical cancer prevention”. In: Frontiers in Applied Mathematics and Statistics 11 (Jan. 28, 2026). ISSN: 2297-4687. DOI: 10.3389/fams.2025.1677512. URL: https://www.frontiersin.org/journals/applied-mathematics-and-statistics/articles/10.3389/fams.2025.1677512/full (visited on 05/02/2026).

[25] Fednant O. Okware, Samuel B. Apima, and Amos O. Wanjara. “Mathematical Modelling of Human Papillomavirus (HPV) Dynamics with Vaccination Incorporating Optimal Control Analysis”. In: Asian Research Journal of Mathematics 19.11 (Oct. 16, 2023), pp. 36–51. ISSN: 2456-477X. DOI: 10.9734/arjom/2023/v19i11751. URL: https://journalarjom.com/index.php/ARJOM/article/view/751 (visited on 05/02/2026).

[26] Shasha Gao et al. “A Dynamic Model to Assess Human Papillomavirus Vaccination Strategies in a Heterosexual Population Combined with Men Who have Sex with Men”. In: Bulletin of Mathematical Biology 83.1 (Jan. 2, 2021), p. 5. ISSN: 1522-9602. DOI: 10.1007/s11538-020-00830-y. URL: https://doi.org/10.1007/s11538-020-00830-y (visited on 05/02/2026).

[27] Carly M. Malburg et al. “Population Size Estimation of Men Who Have Sex With Men in Low- and Middle-Income Countries: Google Trends Analysis”. In: JMIR Public Health and Surveillance 11.1 (Jan. 9, 2025), e58630. DOI: 10.2196/58630. URL: https://publichealth.jmir.org/2025/1/e58630 (visited on 06/18/2026).

[28] Population Pyramids. Nigeria Population Pyramid 2025 - Demographics & Birth Statistics — 16,269 Daily Births. URL: https://www.populationpyramids.org/nigeria (visited on 04/02/2026).

[29] Shauna Stahlman et al. “Online Sex-Seeking among Men Who Have Sex with Men in Nigeria: Implications for Online Intervention”. In: AIDS and behavior 21.11 (Nov. 2017), pp. 3068–3077. ISSN: 1090-7165. DOI: 10.1007/s10461-016-1437-3. URL: https://pmc.ncbi.nlm.nih.gov/articles/PMC5124554/ (visited on 05/03/2026).

[30] Ronel Sewpaul et al. “Initiation, cessation and relapse of tobacco smoking over a 3-year period among participants aged ≥ 15 years in a large longitudinal cohort in rural South Africa”. In: PLOS Global Public Health 5.2 (Feb. 25, 2025), e0004126. DOI: 10.1371/journal.pgph.0004126.

[31] Nigeria Life Expectancy (1950-2025). URL: https://www.macrotrends.net /global-metrics/countries/nga/nigeria/life-expectancy (visited on 04/10/2026).

[32] Daniel Beachler et al. “An examination of HPV16 natural immunity in men who have sex with men (MSM) in the HPV in Men (HIM) study”. In: Cancer epidemiology, biomarkers & prevention : a publication of the American Association for Cancer Research, cosponsored by the American Society of Preventive Oncology 27.4 (Apr. 2018), pp. 496–502. DOI: 10.1158/1055-9965.EPI-17-0853.

[33] Adams Victor Eseoghen et al. “Assessment of the prevalence, regional differences and determinants of tobacco use among men and women of reproductive age in Nigeria”. In: Discover Public Health 22.1 (June 23, 2025), p. 357. DOI: 10.1186/s12982-025-00754-9.

[34] John Doorbar. “The human Papillomavirus twilight zone – Latency, immune control and subclinical infection”. In: Tumour Virus Research 16 (Dec. 1, 2023), p. 200268. DOI: 10.1016/j.tvr.2023.200268.

[35] Hun Jin Kim et al. “Long-Term Outcomes of Chemoradiation for Anal Cancer Patients”. In: Yonsei Medical Journal 54.1 (Jan. 1, 2013), pp. 108–115. DOI: 10.3349/ymj.2013.54.1.108.

[36] Alan G. Nyitray et al. “Incidence, Duration, Persistence, and Factors Associated With High-risk Anal Human Papillomavirus Persistence Among HIV-negative Men Who Have Sex With Men: A Multinational Study”. In: Clinical Infectious Diseases: An Official Publication of the Infectious Diseases Society of America 62.11 (June 1, 2016), pp. 1367–1374. DOI: 10.1093/cid/ciw140.

[37] Pauline Van den Driessche and James Watmough. “Reproduction numbers and sub-threshold endemic equilibria for compartmental models of disease transmission”. In: Mathematical biosciences 180.1-2 (2002), pp. 29–48.

[38] Maia Martcheva. “Methods for deriving necessary and sufficient conditions for backward bifurcation”. In: Journal of biological dynamics 13.1 (2019), pp. 538–566.

[39] S. M. Blower and H. Dowlatabadi. “Sensitivity and Uncertainty Analysis of Complex Models of Disease Transmission: An HIV Model, as an Example”. In: International Statistical Review/Revue Internationale de Statistique 62.2 (1994), pp. 229–243. ISSN: 0306-7734. DOI: 10.2307/1403510. URL: https://www.jstor.org/stable/1403510 (visited on 06/24/2026).

[40] Simeone Marino et al. “A Methodology For Performing Global Uncertainty And Sensitivity Analysis In Systems Biology”. In: Journal of theoretical biology 254.1 (Sept. 7, 2008), pp. 178–196. ISSN: 0022-5193. DOI: 10.1016/j.jtbi.2008.04.011. URL: https://pmc.ncbi.nlm.nih.gov/articles/PMC2570191/ (visited on 06/24/2026).

[41] Safdar S, Ngonghala Cn, and Gumel Ab. “Mathematical assessment of the role of waning and boosting immunity against the BA.1 Omicron variant in the United States”. In: Mathematical biosciences and engineering : MBE 20.1 (Jan. 2023). ISSN: 1551-0018. DOI: 10.3934/mbe.2023009. URL: https://pubmed.ncbi.nlm.nih.gov/36650762/ (visited on 06/25/2026).

[42] Nigeria Immunization Technical Advisory Group (NGITAG). Updated HPV Technical Dossier on Quadrivalent HPV (Gardasil-4valent) 1 -dose Regimen. NGI-TAG. URL: https://www.nitag-resource.org/.

[43] Efficacy of GARDASIL®9 (Human Papillomavirus 9-valent Vaccine, Recombinant). gardasil9-mv. URL: https://www.merckvaccines.com/gardasil9/efficacy/ (visited on 06/24/2026).

[44] Haley E. Randolph and Luis B. Barreiro. “Herd Immunity: Understanding COVID-19”. In: Immunity 52.5 (May 19, 2020), pp. 737–741. ISSN: 1074-7613. DOI: 10.1016/j.immuni.2020.04.012. URL: https://pmc.ncbi.nlm.nih.gov/articles/PMC7236739/ (visited on 06/24/2026).

[45] Nittaya Phanuphak et al. “Anal human papillomavirus infection among Thai men who have sex with men with and without HIV infection: prevalence, incidence, and persistence”. In: Journal of acquired immune deficiency syndromes (1999) 63.4 (Aug. 1, 2013), pp. 472–479. ISSN: 1525-4135. DOI: 10.1097/QAI.0b013e3182918a5a. URL: https://pmc.ncbinlm.nih.gov/articles/PMC3700660/ (visited on 06/24/2026).

[46] Matthew B. Schabath et al. “A prospective analysis of smoking and human papillomavirus (HPV) infection among men in The HPV in Men (HIM) Study”. In: International journal of cancer. Journal international du cancer 134.10 (May 15, 2014), pp. 2448–2457. DOI: 10.1002/ijc.28567.

